# Exercise reshapes cellular immunity toward anti-inflammation and diminishes CCR4 levels on LFA-1α^high^CD4^+^ T cells in multiple sclerosis

**DOI:** 10.64898/2026.11.05.26367365

**Authors:** Sebastian Proschinger, Tiffany Wences Chirino, Leon Hoenen, Sina Trebing, Niklas Joisten, David Walzik, Niklas Daube, Gisa Ellrichmann-Wilms, Shiva Ahmadi, Martin Schneider, Piotr Zadora, Dario L. Frey, Sven G. Meuth, Marc Pawlitzki, Lars Masanneck, Jens Kuhle, Tobias Ruck, Robert Rehmann, Alessa L. Henneberg, Christiane A. Opitz, Philipp Zimmer

**Author notes:** Correspondence: Prof. Philipp Zimmer.

## Abstract

Multiple sclerosis (MS) represents a multifaceted neuroinflammatory disease for which regular exercise is now recommended by many healthcare institutions. Although preclinical studies indicate robust immunomodulatory effects of exercise, evidence from human studies is lacking. In this randomized controlled trial, persons with MS (pwMS) were assigned to three months of high-intensity interval training or a passive control condition, with outcomes assessed at baseline, after intervention, and a four- week follow-up. Additionally, a healthy matched control group was included. Besides exercise-induced sustained increases in physical fitness, systemic inflammation was diminished and the T_reg_-to-T_H_17.1 ratio improved after exercise. Untargeted proteomic analysis of peripheral blood mononuclear cells covering >7000 proteins revealed an exercise-induced decrease in proteins implicated in inflammatory diseases (i.e., MTHFD2). This was supported by inferred transcription factor activity analysis, which indicated enhanced polarization of immune cells toward anti-inflammatory phenotypes at follow-up. Using flow cytometry, we identified an LFA-1α^high^CD4^+^ T-cell subset that was associated with disease biomarkers and exhibited increased CCR4 expression compared to healthy matched subjects which was reduced after exercise. Our findings significantly expand knowledge of how exercise training exerts immediate and sustained immunomodulatory effects in pwMS and thereby strengthens the mechanistic rationale for exercise recommendations made by healthcare professionals.

**One Sentence Summary:** Using multiparametric flow cytometry and state-of-the-art mass spectrometry, we demonstrate that exercise training promotes an anti-inflammatory state at both the systemic and immune-cell proteomic levels and ameliorates dysregulated CCR4 expression on disease-associated LFA-1α^high^ CD4^+^ T cells in multiple sclerosis patients.

## Introduction

Multiple sclerosis (MS) is a progressive neuroinflammatory autoimmune disease that primarily affects females, is commonly diagnosed in early adulthood, and affects more than 2.8 million people worldwide (*1*). Disease progression is driven by localized inflammatory processes within the central nervous system (CNS), leading to continuous neurodegeneration and diverse symptom complexes depending on affected CNS structures. The main concept of disease-modifying therapies (DMTs) is the modulation of peripheral immunity to mitigate inflammation-induced neurodegeneration either by preventing immune cell invasion into the CNS, (un-)targeted cell depletion, or dampening cellular activation (*2,3*).

Besides positive effects of regular exercise on patient-reported outcomes in MS(*4*), a growing body of evidence now supports the application of structured exercise programs to beneficially affect immune cell characteristics implicated in the immunopathology of the disease (*5–7*) without causing adverse effects, which are oftentimes a consequence of DMTs. These observations are consistent with exercise- induced anti-inflammatory effects in healthy subjects and other clinical populations (*8,9*) and are complemented by increasing evidence that levels of physical activity and cardiorespiratory fitness (CRF), which are known to be decreased in persons with MS (*10,11*), are associated with disease progression and neurodegeneration (*12,13*). Especially high-intensity interval training (HIIT) has been shown to be more effective in improving CRF both in healthy subjects and persons with MS(*14,15*), but also regarding the modulation of disease-associated cellular and molecular disease markers (*16,17*).

A main goal of drug therapies in MS is to dampen immune cell trafficking across or their attraction toward the blood-brain-barrier which is achieved by targeting adhesion molecules and chemokine receptors (*18,19*). Although preclinical studies report profound immunomodulatory effects of exercise on immune cell infiltration into CNS and lymphoid organs, cytokine production, and disease progression(*20,21*), there is limited translational evidence for these findings (*22*). Furthermore, most clinical studies investigating exercise-induced effects on the immune system in MS are limited by small sample sizes, short intervention periods, or targeted analytical approaches, which restrict the exploratory potential of resulting data. Just recently, unbiased analyses of alterations within the proteome of circulating immune cells after one training session revealed profound intensity-dependent changes (*23*), providing implications of a significant remodeling of the cellular proteome with functional adaptions in the course of a training intervention over several months.

In this randomized controlled trial (RCT), we comprehensively investigated immediate and sustained effects of a three-month HIIT intervention on circulating immune cells in persons with MS and included a healthy control group matched for age, sex, and body mass index (BMI) for comparison. We characterized MS-relevant T cell subsets and their expression of proteins involved in vascular adhesion and chemotaxis using flow cytometry and further assessed exercise-induced changes within the proteome of peripheral blood mononuclear cells (PBMCs) by mass spectrometry-based profiling of more than 7,000 proteins. Besides a pronounced increase in physical fitness, HIIT induced immediate and sustained changes in composite indices of systemic inflammation, while changes within the PBMC proteome at follow-up indicated a shift of innate and adaptive immune cells toward anti-inflammatory phenotypes and a reduced abundance of proteins linked to chronic inflammatory disease (i.e., MTHFD2). We further identified an LFA-1α^high^CD4^+^ T-cell subset that was associated with disease biomarkers and exhibited increased C-C chemokine receptor type 4 (CCR4) expression compared with healthy controls that declined in the intervention group.

Together, these findings advance our understanding of cellular and molecular mechanisms underlying exercise-induced immunomodulation in persons with MS, thereby strengthening the mechanistic basis for current recommendations of exercise and lifestyle physical activity.

## Results

From a total of 142 subjects that were assessed for eligibility, 75 persons with relapsing-remitting MS were randomized either into the intervention (n=39) or passive control group (n=36) (Fig. 1). The matched healthy subjects (n=29) did not differ from the MS cohort regarding age, BMI, sex, and peak oxygen uptake (V͘O_2peak_) (Table 1), with sex-based subgroup analysis similarly revealing no statistically significant differences (Supplementary Table S1). Peak power output (PPO) was lower in the MS cohort as well as in the male subgroup. In total, seven participants were considered drop-outs (relapse: n=4; change in medication: n=2; other: n=1), and four participants were lost to follow-up as they either did not start the intervention or discontinued the training after one session (Fig. 1A). Additionally, four assessments were missed due to illness of participants (post intervention: n=1; follow-up: n=3). Mean attendance (number of successfully conducted exercise sessions) of all participants who completed the study was 88.73% (± 9.7) with one person dropping below 70% (Supplementary Fig. S1A). During the study period, one adverse event (severe cardiovascular exacerbation) was reported which did not result in drop-out. One participant in the intervention group experienced an episode of known atrioventricular nodal reentry tachycardia (AVNRT) during a training session. The session was therefore interrupted, and the tachycardia ceased spontaneously at rest.

**Fig. 1.**
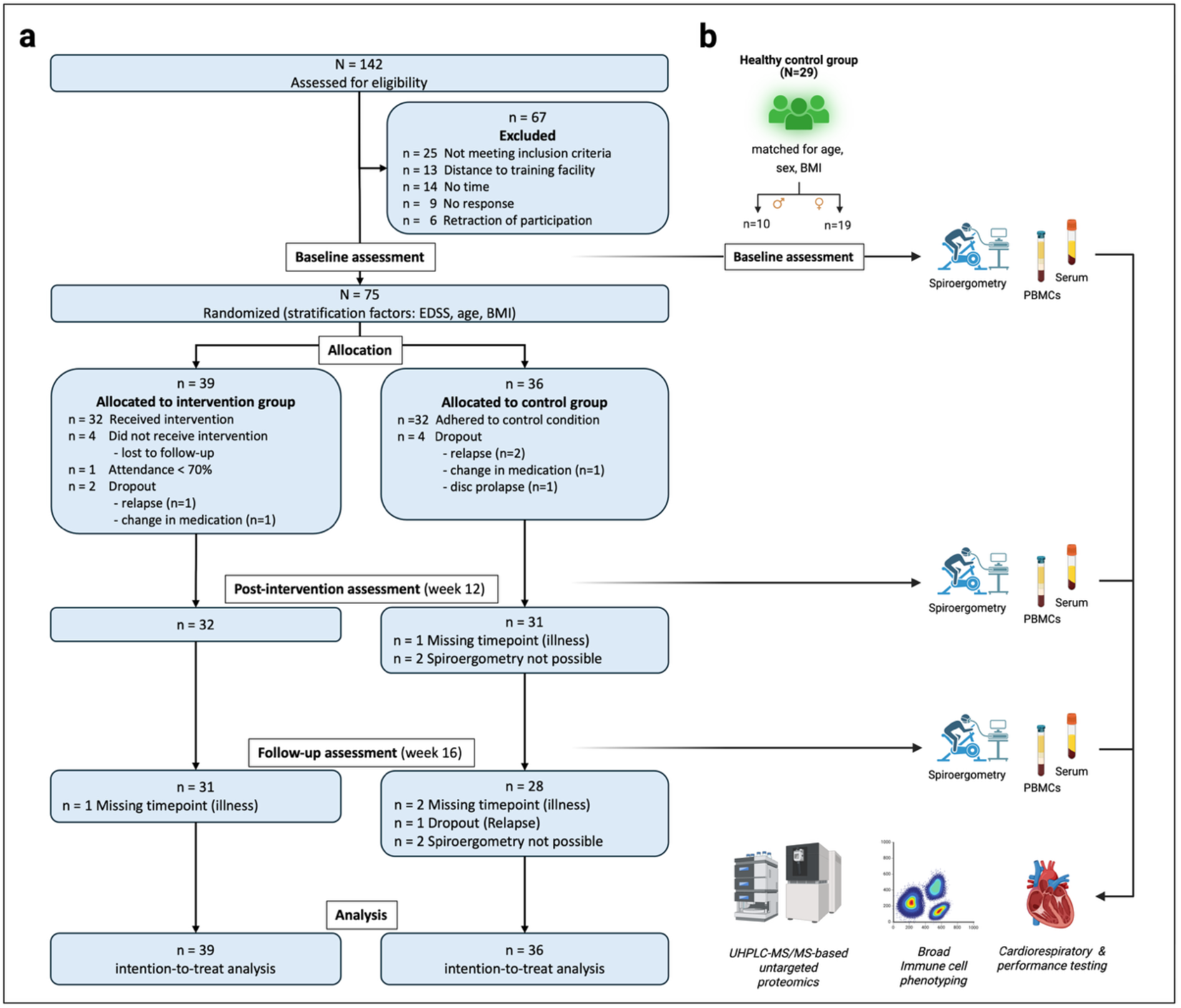
Study design and analysis plan. (a) Flow diagram of the randomized controlled trial according to the CONSORT guidelines (*97*). (b) Additional inclusion of healthy matched subjects, the depiction of assessments per time point, and the analysis of (biomedical) outcomes. BMI: body mass index; EDSS: expanded disability status scale; UHPLC-MS/MS: Ultra-high performance liquid chromatography tandem mass spectrometry; PBMC: peripheral blood mononuclear cells.

**Table 1.** Baseline anthropometric and performance characteristics of included study participants separated by group.

| Characteristic | Intervention group (n=39) | Passive control group (n=36) | Healthy control group (n=29) | p value |
|---|---|---|---|---|
| Sex (m/f) | 12 / 27 | 13 / 23 | 10 /19 |  |
| Age (years) | 41.5 ± 9.5 | 42.5 ± 10.1 | 41.9 ± 11.5 | .96 |
| BMI (kg·m<sup>-2</sup>) | 26.3 ± 5.4 | 25.7 ± 4.5 | 26.1 ± 5.1 | .93 |
| Smoker status (yes / no) | 7 / 32 (22%) | 3 / 33 (9%) | 5 / 24 (21%) |  |
| EDSS | 1.7 ± 1.1 | 1.6 ± 1.3 | n.a. |  |
| Time since diagnosis (years) | 7.5 ± 7.0 | 8.6 ± 8.5 | n.a. |  |
| V̇O<sub>2peak</sub> (mL·kg<sup>-1</sup>·min-<sup>1</sup>) | 30.7 ± 6.1 | 31.6 ± 7.4 | 34 ± 8.4 | .07 |
| PPO (W·kg<sup>-1</sup>) | 2.46 ± 0.58 | 2.53 ± 0.65 | 2.94 ± 0.79 | .003 |
| DMT group |  |  |  |  |
| None | 7 (9.3%) | 5 (6.7%) |  |  |
| Fumaric acid esters | 11 (14.7%) | 8 (10.7%) |  |  |
| Glatiramer acetate | 3 (4%) | 4 (5.3%) |  |  |
| Interferon β | 0 (0%) | 2 (2.7%) |  |  |
| Teriflunomide | 2 (2.7%) | 3 (4%) |  |  |
| Anti-CD20 immunotherapy | 11 (14.7%) | 6 (8%) |  |  |
| Cladribine | 2 (2.7%) | 0 (0%) |  |  |
| S1PR modulators | 3 (4%) | 6 (8%) |  |  |
| Natalizumab | 0 (0%) | 2 (2.7%) |  |  |

Data are presented as mean ± SD. Two-sided unpaired Student´s t-tests were performed to compare the healthy and pooled MS cohort. BMI: body mass index; DMT: Disease-modifying therapy; EDSS: expanded disability status scale; f: female; m: male; PPO: peak power output; S1PR: Sphingosine-1-phosphate receptor; V̇O<sub>2peak</sub>: peak oxygen uptake.

### HIIT induces sustained increases in physical fitness

Consistent with previous studies (*10,11*), we observed that included participants with MS were less physically active compared to matched healthy subjects (p = 4.7×10^-2^, *r_rb_* = .25), as reflected by a lower self-reported metabolic equivalent of task (MET) score, exhibited a lower PPO (p = 2.8×10^-3^, *d* = 0.67) and a trend toward a decreased V͘O_2peak_ (p = 7.0×10^-2^, *d* = 0.40) (Fig. 2A). After the study period, significant interaction effects were shown for V͘O_2peak_ (p = 2.3×10^-9^, ƞp^2^ = .26) and PPO (2.2×10^-16^, ƞp^2^ = .46), with post hoc analyses revealing large increases at post-intervention (V͘O_2peak_: 1.3×10^-11^, *d* = 1.85; PPO: p = 4.1×10^-26^, *d* = 3.30) and follow-up (V͘O_2peak_: 8.2×10^-14^, *d* = 2.09; PPO: p = 4.5×10^-20^, *d* = 2.73) (Fig. 2B). Group differences for both parameters at post- intervention (V͘O_2peak_: 1.9×10^-10^, *d* = 1.93; PPO: p = 1.0×10^-22^, *d* = 3.42) and follow-up (V͘O_2peak_: 1.5×10^-11^, *d* = 2.13; PPO: p = 1.5×10^-16^, *d* = 2.84) were large, although the participants’ reported weekly leisure-time activity returned to baseline levels at follow- up (Supplementary Fig. S1B) underscoring the lasting effects of the HIIT intervention on physical fitness. These results support the notion that HIIT is an effective measure to increase V͘O_2peak_ in persons with MS (*14*) with observable effects lasting for at least four weeks after the end of supervised training. The results further highlight its role to improve and preserve PPO, a measure of the physical functional reserve which is oftentimes neglected in clinical exercise studies but relevant in MS due to a disease- related decline in motor function especially in the lower limbs (*24*). As expected, the increase in V͘O_2peak_ and PPO from pre- to post-intervention positively correlated with the training attendance (Supplementary Fig. S1C), indicating that a regular participation in structured exercise interventions is important to achieve sufficient cardiorespiratory and musculoskeletal adaptations.

**Fig. 2.**
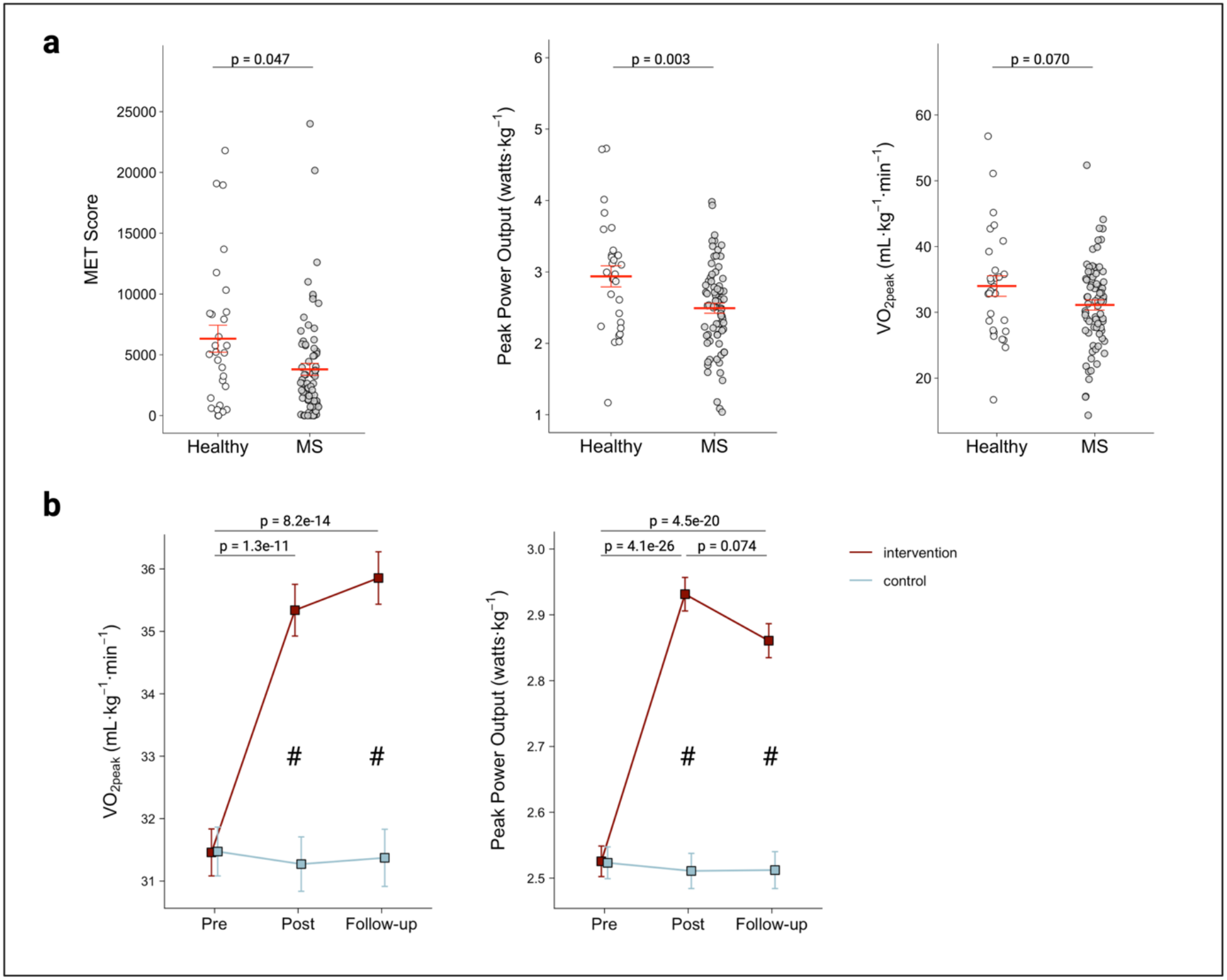
Impact of MS and exercise on physical activity, V͘O_2peak_ and peak power output. (a) Differences in the physical activity level, quantified as self-reported MET score via the Global Physical Activity Questionnaire, and objectively quantified physical fitness parameters between participants with MS and healthy subjects. (b) Exercise- induced improvements in peak power output and V͘O_2peak_. Data are presented as mean ± SEM. ^#^ indicate a significant group difference. MET: Metabolic equivalent of task; MS: Multiple sclerosis, V͘O_2peak_: peak oxygen uptake.

### Modulation of cellular inflammation after HIIT

To improve our understanding of the immunobiological adaptations underlying enhanced physical fitness in persons with MS, we next sought to assess inflammatory activity on a systemic level using the Neutrophil-to-Lymphocyte Ratio (NLR) and the Systemic Immune-inflammation Index (SII) as established inflammatory indices (*25,26*). The MS cohort exhibited a higher NLR (7.2×10^-3^, *r_rb_* = .34) compared to healthy subjects, whereas the SII was not different between both groups (Fig. 3A). When comparing the intervention and control groups over time, a significant group effect was observed for the SII (p = 1.1×10^-2^, ƞp^2^ = .09) with post-hoc analysis revealing lower levels in the intervention group at post-intervention (p = 2.0×10^-2^, *d* = 0.60) and follow- up (p = 2.9×10^-2^, *d* = 0.59). The decrease from pre-intervention to follow-up was moderate (*d* = 0.55) but non-significant (p = 7.7×10^-2^). No significant main effect was observed for the NLR. These results partly confirm previous findings showing a superior effect of HIIT on the SII and NLR in persons with MS compared to moderate continuous exercise (*17*) and extend these findings by a lasting exercise-induced effect on the SII. Interestingly, delta changes in the NLR from pre- to post-intervention were moderately correlated with changes in the V͘O_2peak_ (p = 1.8×10^-2^, *r* = -.41), whereas changes in the SII showed a small but non-significant correlation (p = 1.9×10^-1^, *r* = -.23) (Supplementary Fig. S2) which indicates that the NLR is responsive to changes in the V͘O_2peak_.

**Fig. 3.**
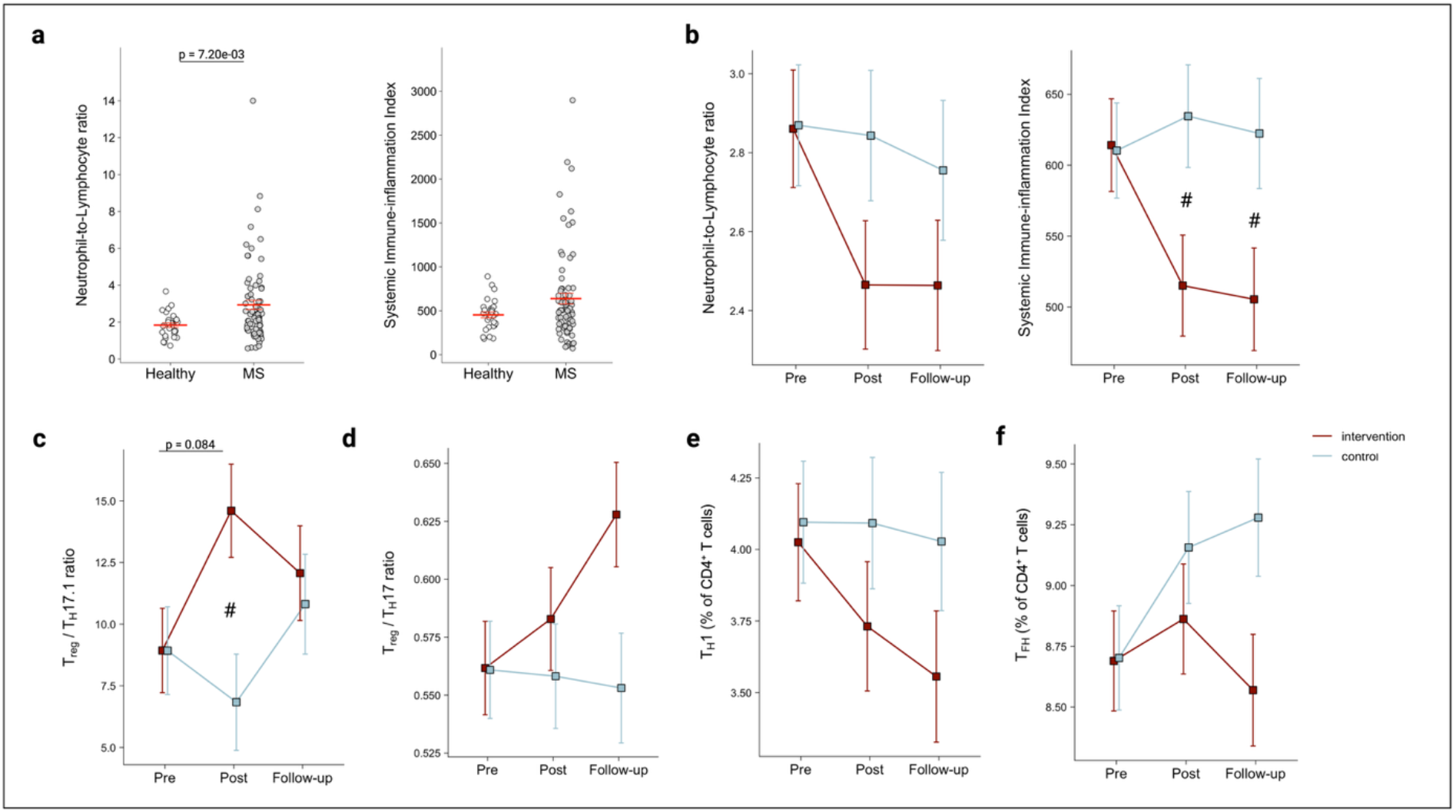
Exercise-induced effects on cellular inflammation indices and T cell subsets implicated in MS. (a) Differences in cellular inflammation indices between participants with MS and healthy subjects. (b) Exercise-induced effects on both indices as well as on (c-f) disease-associated T cell subsets and ratios. Data are presented as mean ± SEM. ^#^ indicate a significant group difference. MS: Multiple Sclerosis; T_H_1: CD4^+^ T helper 1 cells; T_FH_: CD4^+^ follicular helper T cells; T_reg_: CD4^+^ regulatory T cells; T_H_17: CD4^+^ T helper 17 cells; T_H_17.1: CD4^+^ T helper 17.1 cells.

To assess exercise-induced effects on immune cell subsets implicated in disease activity and progression, we next performed flow cytometry analysis of PBMCs (see Supplementary Fig. S3 for gating strategy). Although we did not observe significant main effects for regulatory CD4^+^ T cells (T_regs_), T helper (T_H_) 17, and T_H_17.1 cells (Supplementary Table S2), a significant group effect was shown for the T_reg_-to-T_H_17.1 ratio (p = 4.9×10^-2^, ƞp^2^ = .02) with post hoc analysis revealing a moderate-to-large group difference at post-intervention (p = 4.8×10^-3^, *d* = 0.73) (Fig. 3C). This might be of therapeutic relevance as T_H_17.1 cells are increasingly recognized as a substantial contributor in MS immunopathology (*27,28*), while the induction of T_regs_ is a favorable outcome of therapeutic approaches *(2S)*. Despite no significant main effect was observed for the T_reg_-to-T_H_17 ratio, a moderate increase was shown from pre- intervention to follow-up in the intervention group (*d* = 0.59) as well as a moderate-to- large group difference at follow-up (*d* = 0.66) (Fig. 3D). Similarly, no significant main effects were observed for other CD4^+^ T cell subsets associated with MS (Supplementary Table S2), i.e., T_H_1 and T follicular helper (T_FH_) cells (Fig. 3E,F), but group differences at follow-up were moderate for T_H_1 (*d* = 0.59) and moderate-to-large for T_FH_ (*d* = 0.64).

### HIIT restructures the proteome of circulating immune cells

We next investigated whether exercise-induced changes in cellular inflammation markers were also reflected by alterations in the PBMC proteome. This is of particular relevance since proteomic profiling provides functional insights into cellular states which cannot be captured by phenotypic characterization alone. Using state-of-the-art UHPLC-MS/MS instrumentation, a total of 9,465 proteins were identified and 8,949 proteins were quantified, respectively, across all samples, thereby representing a comprehensive data resource that exceeds results from other studies in the field of immunology and MS (*30,31*).

#### Assessment of data robustness and potential confounders

Quantification of proteins per group and time point (ranging from 7,266 to 7,496, Supplementary Fig. S4A) as well as the inter-individual variability of all quantified proteins (ranging from 28.36% to 32.10%, Supplementary Fig. S4B) were highly consistent. After data preprocessing, the dataset comprised 7,111 proteins that were included into the final analysis. Data imputation across all time points per group showed a well-balanced and homogeneous pattern (Supplementary Fig. S4C), indicating that the imputation procedure preserved the overall data structure without introducing apparent systematic bias. Although the distribution of DMTs among both groups was comparable and remained largely consistent over time (Table 1, Supplementary Table S3), we assessed whether the PBMC proteome of DMT subgroups cluster together due to their well-known effects on circulating immune cells (*2,3*) to rule out any bias arising from medication-induced proteomic alterations. Uniform Manifold Approximation and Projection (UMAP) clustering revealed that participants receiving the same DMT were distributed heterogeneously across the data space and did not form distinct clusters, indicating that DMT-specific effects were not prominently reflected in the PBMC proteome (Supplementary Fig. S4D). Results from flow cytometry analysis (Supplementary Table S2) did not reveal large shifts in major immune cell populations in either group which was complemented by a group-by-group comparison of major immune cell populations per time point (Supplementary Fig. S4E), ruling out that group, time, and group × time interaction effects arise from differences within the PBMC compartment.

#### HIIT induces a coordinated decline in mitochondrial proteins

Using linear mixed model analysis, a total of 34 group × time interaction effects, 1 group effect, and 402 time effects were identified (Fig. 4A). Whereas exercise-induced effects on the proteome at post-intervention were scarce, profound time and interaction effects were observed with a particular focus on changes within the intervention group of 376 differentially abundant proteins. Inspection of significant interaction effects at post-intervention revealed four proteins that were significantly downregulated in the intervention group (Fig. 4B). Among them is PPM1K (p = 3.5×10^-3^, log_2_FC = -0.49), a protein that is predominantly located in the mitochondrial matrix and necessary for branched-chain amino acid metabolism. SNX14 (p = 3.1×10^-2^, log_2_FC = -0.51) is involved in lipid metabolism, mitochondrial transport and anchored to the ER, whereas SEC61G (p = 7.1×10^-3^, log_2_FC = -1.1) plays an important role in transporting polypeptides into the ER for protein folding and maturation. These results suggest that immediate effects of chronic exercise predominantly led to changes in mitochondrial metabolism of non-glucose-derived substrates, but also the translational machinery in immune cells of persons with MS.

**Fig. 4.**
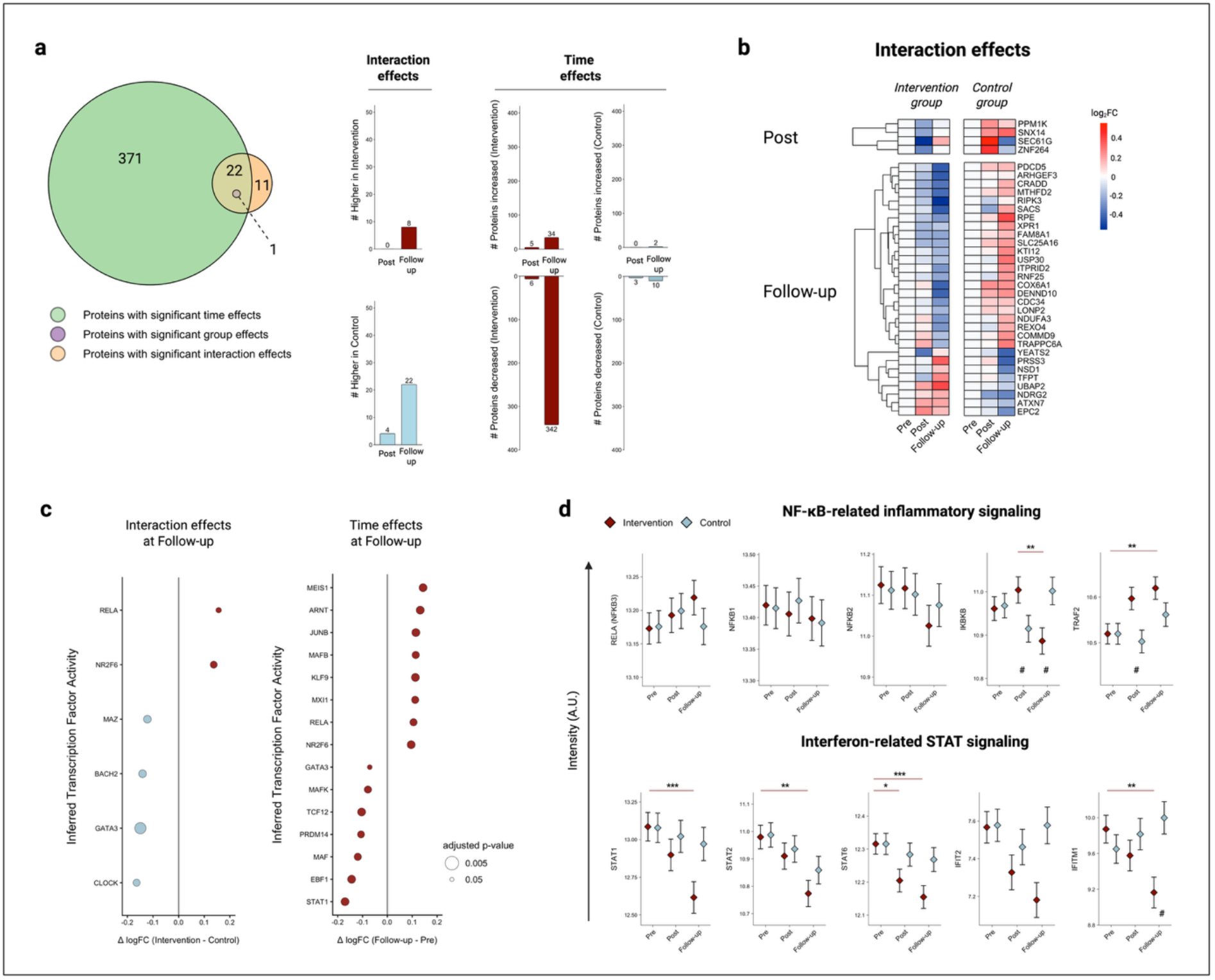
HIIT shifts the PBMC proteome toward a profile consistent with an anti- inflammatory state. (a) Total number of differentially abundant proteins (DAP) with time, group, and group × time interaction effects (left) as well as the total amount of interaction and time effects per group and timepoint (right). (b) Heatmap of interaction effects at post and follow-up including a dendrogram that depicts clusters of proteins with similar kinetics. (c) Inferred changes in transcription factor activity in the intervention group from pre to follow-up (as time effects, left) and between groups at follow-up (as interaction effects, right) based on the abundance of target proteins (n ≥ 20). (d) Targeted analysis of proteins derived from the proteomics dataset that belong to the NF-κB (upper diagram) or STAT (lower diagram) signaling pathway. Data are presented as mean ± SEM in (d). *different from Pre (.05 ≥ p > .005). **different from Pre (.005 > p ≥ .001). ***different from Pre (p ≤ .001). ^#^ indicate a significant group difference.

At follow-up, a total of 30 interaction effects were identified of which 22 proteins were significantly downregulated in the intervention group (Fig. 4B). Although a different set of proteins was identified, a similar picture of altered mitochondrial metabolism could be drawn from the reduced abundances of NDUFA3 (p = 4.1×10^-2^, log_2_FC = -0.52) and COX6A1 (p = 2.7×10^-2^, log_2_FC = -0.74) that belong to mitochondrial respiratory complex I and IV (*32*), and SLC25A16 (p = 1.2×10^-2^, log_2_FC = -0.39) which represents a mitochondrial transporter facilitating the exchange of molecules between the cytosol and the mitochondrial matrix (*33*). Similarly, RIPK3 (p = 3.8×10^-2^, log_2_FC = -0.67) and PDCD5 (p = 2.2×10^-2^, log_2_FC = -0.63) affect mitochondria either by enhancing aerobic respiration and subsequent mitochondrial ROS production (*34*) or by activating the mitochondria-related apoptotic pathway (*35*). Methylenetetrahydrofolate dehydrogenase 2 (MTHFD2) (p = 2.4×10^-2^, log_2_FC = -0.58), located in the mitochondrion and involved in nucleotide synthesis and redox homeostasis, represents a critical protein necessary for activated CD4^+^ T cells to maintain *de novo* purine synthesis(*36*). Sugiura et al. (*36*) further showed that blockage of MTHFD2 promoted T_reg_ and repressed T_H_17 differentiation, and that MTHFD2 is highly upregulated in CNS-infiltrating CD4^+^ T cells in a mouse model of MS which highlights this protein as a potential candidate that drives exercise-induced beneficial effects on CD4^+^ T cells in MS.

#### Anti-inflammatory adaptations of circulating immune cells are mirrored by inferred transcriptional regulation after HIIT

To evaluate the impact of HIIT on transcriptional regulation of PBMCs, we next applied a computational approach to infer transcription factor (TF) activity based on the abundance of proteins encoded by downstream TF target genes within our dataset (*37*). In line with the number of identified time and interaction effects on the protein level (Fig. 4A), pronounced differences in inferred TF activity were identified at follow- up, whereas no significant differences were revealed at post-intervention. Within the passive control group, MAZ (p = 2.4×10^-2^, log_2_FC = 0.092) represented the only TF whose activity was significantly upregulated (Supplementary Fig. S4F). MAZ has been shown to play a significant role in increasing gene expression of STAT1 (*38*) which is essential for interferon signaling and T_H_1 differentiation (*39*). In contrast, the inferred activity of several TFs was modulated in the intervention group over time with the strongest repressive effect observed for STAT1 activity (p = 2.0×10^-2^, log_2_FC = -0.170) (Fig. 4C), thereby contrasting the results observed in the passive control group. An exercise-induced modulatory effect on CD4^+^ T cell differentiation was suggested by a decrease in GATA3 activity (p = 4.4×10^-2^, log_2_FC = -0.071), the master transcription factor of T_H_2 cells, or an increase in NR2F6 activity (p = 2.0×10^-2^, log_2_FC = 0.096) which has been shown to counteract T_FH_ cell differentiation (*40*). The increased activity of JunB (p = 2.0×10^-2^, log_2_FC = 0.115) can be linked to enhanced T_reg_ differentiation (*41,42*), although experimental evidence from other studies indicate T_H_17-promoting effects (*43*), suggesting context-dependent effects that determine downstream JunB signaling. Similarly, an increased ARNT activity (p = 2.0×10^-2^, log_2_FC = 0.132) is indicative of an enhanced CD4^+^ regulatory cell type- or T_H_17- signature depending on whether the complexing partner of ARNT is the aryl hydrocarbon receptor (*44,45*) or hypoxia-inducible factor 1-α (*46*).

Regarding effects on myeloid cells, increased MEIS1 activity (p = 2.0×10^-2^, log_2_FC = 0.144) indicates an enhanced differentiation of anti-inflammatory M2 macrophages (*47*) or monocytic myeloid-derived suppressor cells (*48*). The increases in KLF9 (p = 2.0×10^-2^, log_2_FC = 0.113) and MAFB activity (p = 2.4×10^-2^, log_2_FC = 0.114) were consistent with the aforementioned observations as KLF9 inhibits STAT1 transcription and enhances M2 macrophage polarization (*49*), while MAFB contributes to an anti- inflammatory profile of monocyte-derived macrophages (*50*). On the other hand, an increase in RELA activity (p = 2.4×10^-2^, log_2_FC = 0.105), a canonical NF-κB subunit and important for T_H_17 polarization and cytokine production in MS (*51*), implies an adverse effect on immune cell signaling in response to exercise. Targeted analysis of proteins involved in the NF-κB signaling pathways showed that the expression of RELA itself and other relevant proteins (i.e., NFKB1, NFKB2) remained mainly unchanged over time and between groups (Fig. 4D, Supplementary Fig. S5A). In contrast, the abundance of STAT1 significantly decreased from pre-intervention to follow-up in the intervention group and was complemented by similar kinetics for STAT2, STAT6, and other related proteins (Fig. 4D, Supplementary Fig. S5B) which underlines the finding of a decrease in inferred STAT1 activity. Interestingly, CLOCK activity was significantly reduced at follow-up in the intervention group compared to the passive control group (p = 2.6×10^-2^, log_2_FC = -0.164) (Fig. 4C). Besides being a core component of the transcriptional circadian clock machinery, CLOCK is implicated in the activation of NF- κB (*52*) and the NLRP3 inflammasome (*53*) which contrasts the finding of increased RELA activity (p = 3.7×10^-2^, log_2_FC = 0.156) in the intervention group. Although enhanced NR2F6 (p = 2.6×10^-2^, log_2_FC = 0.137) and decreased MAZ (p = 2.1×10^-2^, log_2_FC = -0.122) activities in the intervention group compared to the passive control group indicate a reduced STAT1 signaling and an attenuated induction of T_FH_ differentiation (*38,40*), a decreased BACH2 activity (p = 2.1×10^-2^, log_2_FC = -0.141) implicates a diminished ability to stabilize T_reg_ function and to repress T_H_17-mediated inflammatory signaling (*54,55*).

### CCR4 expression is increased on disease-associated LFA-1α^high^ CD4^+^ T cells in MS and reduced following exercise

In addition to reducing peripheral inflammation, DMTs further limit immune cell recruitment to and migration across CNS barriers by modulating chemokine receptors and adhesion molecules (*2,3,18*). Besides very late antigen-4 (VLA-4), the lymphocyte function-associated antigen 1 (LFA-1) represents another disease-associated adhesion molecule whose expression on lymphocytes is higher in persons with MS (*56*). Reducing or abrogating LFA-1 expression on peripheral CD4^+^ T cells in mice significantly hampered their CNS-invading potential and reduced neuroinflammation (*57,58*), whereas blockage of the alpha chain of LFA-1, namely LFA-1α, similarly diminished transmigration of activated CD4^+^ T cells from persons with MS (*59*). Here we observed that CD4^+^ T cells expressing LFA-1α can be discriminated into an LFA- 1α^low^ and an LFA-1α^high^ population (Fig. 5A). To elucidate whether both populations can be discriminated by their cumulative potential for migration and chemoattraction, we next analyzed LFA-1α^low^ and LFA-1α^high^ subsets for their protein expression of other adhesion molecules (i.e. PSGL-1 and CD49d) and chemokine receptors (i.e., CCR4, CCR6, CXCR3, CXCR5, CXCR6, and CX_3_CR1). We observed that the LFA-1α^high^ CD4^+^ subset expressed a significantly higher cumulative amount of both adhesion molecules and chemokine receptors (Fig. 5B). Similar results were observed for CD8^+^LFA-1α cells (Supplementary Fig. S6A). Given the central role of immune-cell recruitment and migration in MS immunopathology, we examined whether the frequencies of LFA-1α^high^ CD4^+^/CD8^+^ subsets were associated with disease biomarkers and clinical characteristics. Indeed, significant associations were observed with serum glial fibrillary acidic protein (sGFAP), serum neurofilament light chain (sNfL), NLR, and disease duration (Fig. 5C), thereby corroborating a potential pathogenic role of LFA-1α^high^ T cells in MS. However, no significant exercise-induced changes in these parameters were observed (Supplementary Table S2). Based on these disease-associated characteristics of LFA-1α^high^ T cells, we were further interested whether a dysregulated, disease-specific signature of certain chemokine receptors or adhesion molecules on these subsets exist. A detailed analysis of chemokine receptor and adhesion molecule expression within LFA-1α^high^ subsets compared to healthy subjects was conducted and revealed significantly higher levels of CCR4 (p = 1.2×10^-4^, *r_rb_* = .49) and PSGL-1 (p = 1.9×10^-4^, *r_rb_* = .47) on LFA-1α^high^ CD4^+^ cells, and CX_3_CR1 (p = 1.9×10^-2^, *r_rb_* = .30) on CD8^+^LFA-1α^high^ cells (Fig. 5D-F, Supplementary Fig. S6B). In regard to exercise, a significant group effect (p = 1.1×10^-2^, ƞp^2^ = .09) was observed for CCR4 expression, showing a decrease from baseline to follow-up (p = 3.7×10^-3^, *d* = .80) and significant group differences both at post- intervention (p = 4.9×10^-3^, *d* = .77) and follow-up (p = 5.0×10^-2^, *d* = .55) (Fig. 5D). These results indicate a beneficial exercise-induced effect on the chemoattractive potential of this disease-associated LFA-1α^high^ CD4^+^ T cell subset, as both LFA-1 and CCR4 are increasingly investigated in MS and CNS inflammation (*57–61*).

**Fig. 5.**
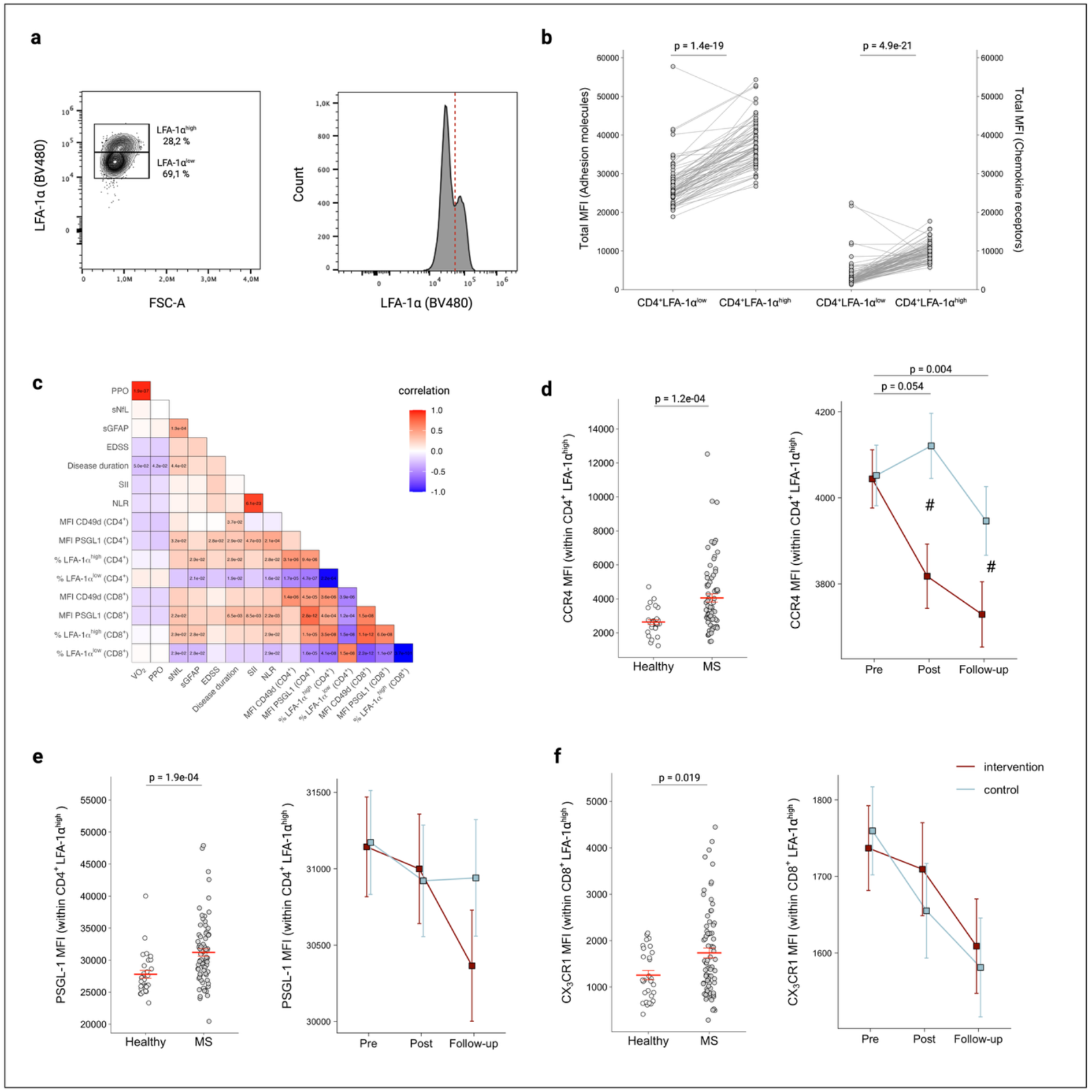
Identification of disease-associated LFA-1α^high^ CD4^+^ T cells and reduction of dysregulated CCR4 expression following HIIT. (a) Discrimination of CD4^+^LFA- 1α^+^ T cells into LFA-1α^high^ and LFA-1α^low^ cells via dot plot and histogram visualization (see also Supplementary Fig. S3 for CD8^+^ T cells). (b) Higher expression of cumulative adhesion molecule (left) and chemokine receptor expression (right) on LFA-1α^high^ CD4^+^cells compared to CD4^+^LFA-1α^low^ cells. (c) Correlation heatmap including MS-related clinical measures, biomarkers, and CD4^+^/CD8^+^ T cell-derived characteristics. For sGFAP and sNfL, their z-score was used. (d-f) Expression levels of chemokine receptors and adhesion molecules within the LFA-1α^high^ subset of CD4^+^/CD8^+^ T cells that are significantly different between MS and healthy subjects as well as exercise- induced effects on their protein expression. Data are presented as mean ± SEM. ^#^ indicate a significant group difference. sGFAP: serum glial fibrillary acidic protein; MFI: Mean fluorescence intensity; MS: Multiple sclerosis; sNfL: serum neurofilament light chain; NLR: Neutrophil-to-Lymphocyte ratio; PPO: Peak power output; SII: Systemic immune-inflammation index; V͘O_2_: peak oxygen uptake.

## Discussion

Treatment of MS necessitates continuous drug treatment targeting peripheral immune cells to decelerate disease progression. Although increasing evidence supports immunomodulatory effects of exercise in MS on the cellular level, a comprehensive analysis from a well-characterized randomized trial with a robust study design is lacking. In this randomized controlled trial, we assessed immediate and lasting effects of HIIT over three months on the peripheral immune system in persons with MS. This was complemented by the inclusion of an age-, BMI-, and sex-matched healthy control group that was used for validation and strengthening of findings. Using state-of-the-art mass spectrometry, this RCT is the first to assess alterations within the immune cell proteome of persons with MS following chronic exercise. Besides profound and sustained increases in physical fitness, we found peripheral adaptations on the cellular and molecular level that indicates a shift toward an increased anti-inflammatory state. Flow cytometry analysis further identified a disease-associated LFA-1α^high^ CD4^+^ T cell subset with increased surface expression of CCR4 compared to healthy subjects that was reduced after exercise. These results underscore the effectiveness of HIIT in persons with MS and significantly expand the knowledge of how exercise training exerts immunomodulatory effects in MS with further implications for future studies.

The observed improvements in V͘O_2peak_ are in line with other studies in the field (*14*) and indicate, together with the low drop-out rate and high training attendance, that our training program was well-accepted and effective. We further showed that the increase in V͘O_2peak_ was sustained at least for four weeks, although the participants’ reported weekly leisure-time activity returned to baseline levels at follow-up. This rules out a potential bias arising from a continuation of exercise after the intervention period. The profound increase in PPO should be emphasized, since this measure is not reported frequently (*14*) but constitutes a relevant feature in MS due to the disease-related decline in motor function especially in the lower limbs (*24*). Although not significant, a trend (p = .074) for a decline in PPO from post-intervention to follow-up was observed, whereas the V͘O_2peak_ remained unchanged, suggesting more sustainable adaptations of the cardiorespiratory system compared to the musculoskeletal system. So far, only one pilot study assessed the effects of exercise training on skeletal muscle morphology and metabolism in MS without including a follow-up time point (*62*), so that future studies may elaborate on these observations.

The diminishing effects of HIIT on the SII remained stable until follow-up which confirms and extends previous findings from another RCT involving persons with MS (*17*). On the subset level, we showed that the T_reg_-to-T_H_17.1 ratio was significantly higher in the intervention group at post-intervention which did not sustain until follow- up, indicating a beneficial but temporary effect that necessitates continuous exercise to persist. Although regular exercise has been shown to be a promising strategy to modulate T_H_17-related outcomes (*6,7*), we were the first to assess exercise-induced changes on T_H_17.1 cells which are increasingly discussed in MS immunopathology (*27,28*).

Using state-of-the-art UHPLC-MS/MS-based analysis of the PBMC proteome, we identified considerably more differentially abundant proteins at follow-up compared to post-intervention with most of them being decreased in the intervention group. This suggests that a recovery period with reduced cardiovascular and musculoskeletal strain may be necessary for (delayed) proteomic and immunometabolic adaptations to emerge in circulating immune cells in response to training. Our findings point to an altered mitochondrial metabolism indicative of a diminished mitochondrial respiration or a reduced synthesis of macromolecules, lipids, and nucleotides that leads to an attenuated proliferation of immune cells (*63*), a mode of action of some DMTs (e.g., teriflunomide or cladribine). The reduced abundance of MTHFD2 in the intervention group is particularly relevant in the context of MS, as MTHFD2 downregulation limited the proliferation and migration of autoreactive CD4^+^ T cells while promoting T_reg_ differentiation (*36*). Indeed, reducing MTHFD2 activity has just recently been proposed as a therapeutic strategy to suppress sustained T-cell-mediated inflammation in rheumatoid arthritis (*64*). Therefore, future studies in this field may focus on MTHFD2 as a potential candidate of exercise-induced effects on CD4^+^ T cell metabolism and differentiation.

In line with the observations on the protein level, the majority of alterations in inferred TF activity point toward gene regulation that favored an anti-inflammatory state which is primarily supported by a decrease in inferred STAT1 activity and increases in NR2F6, MAFB, KLF9, and MEIS1 activity, respectively. While STAT1 transcription is necessary for T_H_1 differentiation (*39*), an increased activity of NR2F6 is capable of inhibiting T_FH_ cell differentiation (*40*), cell types that are implicated in the immunopathology of MS (2). Additionally, the inferred increase in MEIS1, MAFB, and KLF9 activity indicate an enhanced polarization of monocytes and macrophages toward an anti-inflammatory state (*47–49*). Monocytes are largely overlooked in the context of exercise-induced effects in MS despite a growing attention in recent years (*65*), so that future studies may focus on the effects of regular exercise on circulating monocytes. The interpretation of the inferred increases in both JunB and ARNT activity is less clear, since experimental evidence exists for their induction and stabilization of the T_reg_ (*41,42,44,45*) or T_H_17 lineage (*43,46*), respectively. Although exercise-induced activation of the aryl hydrocarbon receptor in PBMCs has been reported (*66*), future studies should characterize both upstream regulators and downstream signaling of JunB and ARNT in more detail. Results from the observed interaction effects of inferred TF activities support an increase in NR2F6 and a decrease in MAZ in the intervention group complemented by a decrease in CLOCK activity, indicating decreased inflammatory signaling and a diminished potential of T_FH_ cell differentiation (*38,40,52,53*). However, the altered activities of RELA and BACH2 contrast these findings by their involvement in NF-κB signaling (RELA) and immunoregulatory and T_reg_-promoting characteristics in CD4^+^ T cells (BACH2) (*51,54,55*). Taken together, most changes in the PBMC proteome indicate a shift toward an anti-inflammatory state both on the protein and inferred TF activity level, although some results suggest a more complex or partially divergent response. Since the results on transcriptional regulation were based on inferential TF activity modelling and reflect changes in the abundance of proteins encoded by downstream TF target genes, future studies should elaborate on these findings by validating an activation or inhibition of highlighted TF as well as associated signaling pathways.

Using flow cytometry, we further identified an LFA-1α^high^ T cell subset that exhibited a significantly higher cumulative expression of other adhesion molecules and chemokine receptors compared to the LFA-1α^low^ subset and is associated with disease biomarkers and clinical measures of MS. LFA-1α represents the larger subunit of the heterodimer LFA-1, is almost exclusively expressed in its dimerized form on the cell membrane (*67*), and is the primary target for modulating the biological function of LFA-1 (*59,68,69*). Findings that attenuation and abrogation of LFA-1 expression on CD4⁺ T cells dampened neuroinflammation by reducing transmigration into the CNS (*57,58*) increase the interest to target LFA-1-expressing cells therapeutically. We found that CCR4 expression on disease-associated LFA-1α^high^ CD4^+^ cells was higher in persons with MS compared to matched healthy subjects and that HIIT significantly diminished CCR4 expression. Indeed, CCR4 is increasingly implicated in inflammatory diseases (*70*), has been identified as a susceptibility gene for MS (*71*), and is constitutively expressed on T_H_17 and T_H_2 cells. It was shown that CCR4^+^ cells are readily able to enter the CNS (*72*) and that CCL22, a cognate ligand for CCR4, is increased in cerebrospinal fluid of persons with MS (*73,74*), leading to an enhanced attraction of peripheral CCR4^+^ cells. In the context of experimental MS, evidence supports the role of CCL17, another cognate ligand of CCR4, in attracting T_H_17 cells to the CNS (*61*), while inhibition of CCR4 signaling hampered T_H_17 and T_H_1 polarization and ameliorated disease progression (*75*). However, no previous study has specifically investigated CCR4⁺ CD4⁺ T cells expressing LFA-1, particularly the LFA-1α^high^ subset, in MS. Our results highlight regular exercise as a strategy to dampen dysregulated CCR4 on a disease-associated cell type expressing high levels of LFA-1α.

Some limitations of this study should be addressed. Based on the exploratory nature of including high dimensional data from untargeted proteomics and multiparametric flow cytometry, the majority of the results reported here were derived from exploratory analyses and should therefore be interpreted accordingly. In addition, the generalizability of the present findings may be limited by the characteristics of the study cohort. Although HIIT over three months was well tolerated and effective in our study cohort, this exercise regimen might not be equally applicable to persons with greater disease severity (EDSS > 4) due to more pronounced motor impairments. Further, our access to biological samples was constrained to the peripheral blood compartment which limits clinically meaningful interpretation in this neuroinflammatory disease. Future studies in this field should therefore acquire cerebrospinal fluid to better translate exercise-induced effects observed in the periphery to potential changes within the CNS. Additionally, we did not perform functional assays in this study (e.g., transmigration assay across human brain endothelial cells, *in-vitro* stimulation assays using MS-specific autoantigens) which would provide mechanistic evidence for our observed findings. Although we assessed exercise-induced changes of thousands of proteins within circulating PBMCs, they are composed of a heterogeneous mixture of adaptive and innate immune cells which mask the identification of cell type-specific alterations. This limitation should be addressed in future studies by sorting cell types of interest such as CD4^+^ T cells and monocytes, thereby validating and extending our findings. Another limitation that needs to be addressed is the heterogeneous use of MS-specific medications within the study cohort, which, although largely stable between groups over the study period, potentially introduced confounding effects.

In conclusion, we substantiate existing evidence on the beneficial effects of HIIT on physical fitness and systemic inflammation in persons with MS and significantly expand this knowledge by our findings on its ability to reshape cellular immunity toward anti- inflammation on the phenotypic and proteomic level. The finding that the strongest changes within the PBMC proteome emerged at follow-up is original and opens an interesting and overlooked perspective on delayed immunometabolic adaptations to training. The observation of an exercise-induced decline in dysregulated CCR4 expression on disease-associated LFA-1α^high^ CD4^+^ T cells proposes another potential mechanism of how exercise exerts immunomodulatory effects in MS. Overall, our results strengthen national and international recommendations on regular exercise for persons with MS by providing biological underpinnings of exercise-induced immunomodulatory effects within the cellular immune system without causing any adverse effects.

## Materials and methods

### Subjects

A total of 75 persons with relapsing-remitting MS participated in this RCT. Subjects were included if they were between 18 and 60 years of age, had approved relapsing- remitting MS, an EDSS between 0 and 4, and approved sports suitability assessed by a medical doctor via an electrocardiogram (ECG) at rest and during exercise (custo diagnostic, custo med GmbH, Ottobrunn, Germany). Study inclusion was further only considered if no adjustments in DMT were made within three months before study commencement. Subjects were excluded if they had any previous history of internal, cardiovascular or orthopedic disease that prohibit study participation, pregnancy or breastfeeding, and if they experienced a relapse or significant worsening of MS-related symptoms during the study period. If a change in DMT was made during the study period, respective time points were not considered for follow-up analysis.

The study was reviewed and approved by the ethics committee of the Leibniz Institute at TU Dortmund University (No. 220, date of approval: June 2022). All subjects signed a written informed consent prior to participation. The study has been prospectively registered at the German Clinical Trials Register (DRKS00033055; https://drks.de/search/en/trial/DRKS00033055; registered 16 November 2023).

Participant enrollment took place from 03 January 2024 (first participant) to 15 October 2025 (last participant).

### Assessments

Participants attended four visits including a baseline testing (week 0), a mid-term visit in week 6, and visits in week 13 (post intervention) and week 17 (post follow-up). At the first visit, written informed consent and disease-related data were obtained which was followed by baseline testing. Each visit took place between 7 a.m. and 11 a.m. at the sports medical laboratory at TU Dortmund University. Participants were asked to arrive fasted and well hydrated, to avoid strenuous exercise (at least 48 hours) and to not drink alcohol, coffee or to take dietary supplements (at least 24 hours) prior to the visit. Questionnaires were filled out online using a link that was provided to the participants 24 hours before each testing day at the TU Dortmund University.

Each visit included the recording of anthropometric data, a blood draw, and cardiopulmonary exercise testing (CPET) until voluntary exhaustion which was conducted on a cycle ergometer (Cortex Bike M) using the MetaLyzer 3B-R2 (CORTEX Biophysik, Leipzig, Germany) during which heart rate was continuously measured (Polar H10). The rating of perceived exertion (RPE, Borg scale (6–20)) (*76*) was recorded every three minutes (min) and immediately after exercise cessation. The highest 15 seconds interval during the CPET was used to calculate V͘O_2peak_ and PPO values. In brief, male and female participants conducted a steep ramp test after a calibration phase of 1 min and a warm-up phase of 2 min at 25 watts (W) for females and 50 W for males, respectively. Based on the participants BMI and the metabolic equivalent score, retrieved from the Global Physical Activity Questionnaire (GPAQ) questionnaire (*77*), a ramp protocol was chosen to increase either 10 or 15 W/min for females and 15 or 20 W/min for males, respectively. Physical activity and sleep behavior was monitored for seven days by a wearable device (ActiGraph^TM^, Pensacola, USA) after baseline testing and before study commencement (week 0), in week 6 of the intervention phase, and in the weeks after the intervention phase (week 13) and follow-up (week 17), respectively.

### Randomization

Using concealed allocation, study participants were randomly assigned into the intervention or passive control group immediately after baseline testing using stratified permuted block randomization with a 1:1 allocation ratio and randomly varying block sizes of four and six. The random allocation sequence was computer-generated in Microsoft Excel by an investigator who was not involved in participant enrollment or assessments. To ensure a balanced group allocation into each study arm, stratification factors were used and dichotomized as follows: age (18-34 vs. 35-60 years), EDSS (0- 2.0 vs. 2.5-4.0), and BMI (< 25 vs. ≥ 25 kg/m^2^).

### Blinding

Due to the nature of the trial, participants could not be blinded to group allocation. However, assessors taking outcome measures after the intervention phase and at follow-up were blinded to group allocation by avoiding any contact between participants and assessors throughout the study period (except for assessments).

### Intervention

The exercise intervention was conducted at the training facility at the TU Dortmund University. Participants allocated to the HIIT group exercised three times per week over a three-month intervention period on a bicycle ergometer (ergoselect5, ergoline GmbH, Germany). This intervention period was split into four blocks of three weeks with progressing intensity from block one to four. The exercise intensity was determined by peak power output (PPO) in watts (W) that was achieved during the baseline assessment. Each training session started with a five min warm-up, comprised a total of seven intervals of 90 seconds that were interspersed by active recovery periods of 3 min between each interval, and ended with a five min cool-down, resulting in a total training duration of 38,5 min. Whereas the intensity during the active recovery phases remained constant over the entire study period (20-25% PPO), the exercise intensity progressively increased from block one to four (block 1: 70-75% PPO; block 2: 80-85% PPO; block 3: 85-90% PPO; block 4: >90% PPO). Since the body adapts to the exercise stimulus over time, another CPET was conducted in intervention week six in order to update the PPO that was used for defining exercise intensity. A detailed overview of the training intervention is provided in Supplementary Table S4. During each training session, the heart rate was continuously measured (ers2 ECG Transmitter, ergoline GmbH, Germany) and the RPE (*76*) was quantified at training start, after the 4^th^ and the 7^th^ interval, respectively. Severe adverse events, defined as i) severe cardiovascular, neurological, orthopedic, or pulmonary diseases (e.g. renal failure, hepatic dysfunction, cardiovascular disease) or ii) severe cardiovascular exacerbations, were recorded during the study period. After finishing the intervention phase, a four-week follow-up period was included into this study in which participants were asked to continue their daily life.

For ethical reasons and to provide all participants the opportunity to benefit from the exercise program, the participants in the passive control group were included in a waiting list to be offered the same exercise program after the study period. Furthermore, all participants were asked not to change their behavior in daily life routines such as exercise, nutrition, or sleep. Participants attending less than 25 training sessions (corresponding to 70% of overall attendance) were considered non-compliant to the study protocol and were therefore excluded from follow-up analysis (*6*). The CPET in week six was considered equivalent to a training session, since the conduction of three training sessions and a CPET within one week was not feasible.

### Healthy control group

Healthy subjects were recruited through public announcements and carefully selected to match participants from the MS cohort for age, BMI and sex. Subjects were included if they were between 18 and 60 years of age, had no internal, neurological or orthopedic diseases, no chronic intake of immunoregulatory or anti-inflammatory drugs, and no illness within four weeks before study participation. The participants were asked to attend one visit at the sports medicine lab which included, similar to the assessment of the MS subjects, the collection of anthropometric data, a blood draw, and a CPET. The ramp protocol was chosen according to a recently published study by Wenzel et al. (*78*). Questionnaires were filled out online using a link that was provided to the participants 24 hours before the testing day at the TU Dortmund University. Written informed consent was obtained prior to participation.

### Blood draw and sample preparation

Blood sampling was performed at each visit by vein puncture after the BIA when participants rested for ten min in a lying position. PBMC purification was done immediately after blood draw from 27mL whole blood, collected in lithium heparin S- Monovettes^®^ (Sarstedt), by density gradient centrifugation at RT using SepMate™ PBMC Isolation Tubes (STEMCELL Technologies). Purified PBMCs were resuspended in 1mL freezing medium (90% FBS, 10% DMSO), stored overnight in a cell freezing container at -80°C, and transferred to -150°C on the next day. For serum isolation, Serum-Gel S-Monovettes^®^ (Sarstedt) were kept in upright position at 4-8°C for 30 min. After clotting, tubes were centrifuged for 10 min at 2500xg at 4°C, aliquoted, and stored at -80°C until analysis.

The same sample preparation procedures were carried out for blood derived from the healthy control subjects.

### Flow cytometry

Flow cytometry analysis was performed using a Cytek® Northern Lights™ full spectrum flow cytometer (Cytek Biosciences, California, USA). Cryopreserved PBMCs were gently thawed in a water bath at 37°C with a mean recovery across all groups and time points of 90.5 ± 4.8% viable cells assessed with the Zombie NIR™ Fixable Viability Kit (BioLegend, San Diego, CA, USA). In brief, 1x10^6^ PBMCs were incubated with a master mix containing Brilliant Stain buffer (BD Biosciences) and antibodies against surface antigens for 30 min at 4°C in the dark. The antibody cocktail comprised anti-CD20 (BV421, clone 2H7), anti-CD3 (Alexa Fluor 700, clone SK7), anti-CD45RA (BV570, clone HI100), anti-CD49d (BV510, clone 9F10), anti-CD62L (BV711, clone DREG-56), anti-CD69 (BV605, clone FN50), anti-CD279 (APC, clone NAT105), anti- CX3CR1 (PE-CF594, clone 2A9-1), anti-CD196 (BV785, clone G034E3), anti-CD194

(PerCP/Fire 806, clone L291H4), anti-CD183 (APC/Fire 810, clone G025H7), anti- CD165 (APC/Fire 750, clone KPL-1) (all from BioLegend, San Diego, CA, USA), anti- CD4 (PerCP, clone SK3), anti-CD8 (BV750, clone SK1), anti-CD25 (BB700, clone M- A251), anti-LFA-1α (BV480, clone G43-25B), anti-CD185 (BB515, clone RF8B2), anti- CD186 (RB744, clone 13B 1E5) (all from BD Biosciences, NJ, USA) and anti-CD56 (violetFluor450, clone MY31) (Cytek Biosciences). After washing, cells were fixed for 40 min at 4°C in the dark using the BD Pharmingen™ Fixation Buffer and then incubated with BD Pharmingen™ permeabilization buffer and anti-Ki-67 (Alexa Fluor 647, clone Ki-67), and anti-Foxp3 (PE, clone 259D/C7) for intracellular staining for 45 min at 4°C in the dark. Cells were acquired on the flow cytometer within two hours after finishing the staining protocol. Gating was performed using FlowJo™ 10.10.1 (see Supplementary Fig. S3 for gating strategy). T_regs_ were phenotyped as CD4^+^Foxp3^+^CD25^high^, T_H_1 cells as CD4^+^CXCR3^+^CCR4^-^CCR6^-^, T_H_2 cells as CD4^+^CXCR3^-^CCR4^+^CCR6^-^, T_H_17 cells as CD4^+^CXCR3^-^CCR4^+^CCR6^+^, T_H_17.1 cells as CD4^+^CXCR3^+^CCR4^-^CCR6^+^, and T_FH_ cells as CD4^+^CXCR5^+^. Monocytes were identified based on SSC and FSC parameters. Mean fluorescence intensity (MFI), quantified as the geometric mean, was calculated in FlowJo™ and used as a relative surrogate marker of the celĺs protein expression level.

### Haemogram

Within 10 min after blood draw into lithium heparin S-Monovettes^®^ (Sarstedt), the absolute cell numbers of major blood cell populations were quantified on a Hematology Analyzer (SYSMEX XP3000, Norderstedt, Germany). The SII was calculated as *(platelets × neutrophils) × lymphocytes^−1^* and the NLR as *neutrophils × lymphocytes^−1^*.

### Serum neurofilament light chain (sNfL) and serum glial fibrillary acidic protein (sGFAP) analysis

Serum was used to measure both sNfL and sGFAP as RUO immunoassays (ECLIA, Electrochemiluminescence immunoassay, two-incubation step sandwich assay) according to the manufacturer’s instructions using the Roche Cobas 8000 (Roche Elecsys^®^ module). All laboratory technicians performing the analyses were blinded to the clinical data and group allocation. Using Generalized Additive Models for Location, Scale and Shape (GAMLSS) from large reference databases (*79,80*), z-scores from the quantified concentration of both parameters by adjusting for age and BMI (sNfL) or for age, BMI, and sex (sGFAP), respectively.

### UHPLC-MS/MS-based untargeted proteomics of PBMCs

#### Sample preparation

In brief, cryopreserved PBMCs were gently thawed in a water bath at 37°C, washed twice with PBS containing 1mM EDTA, and lysed in a commercial RIPA buffer containing 0.1% SDS (Thermo Fisher Scientific, USA), 10mM sodium fluoride (Sigma- Aldrich, Merck, Germany), 1mM sodium orthovanadate (Sigma-Aldrich, Merck, Germany), cOmplete™ Protease Inhibitor Cocktail, PhosSTOP™ (both Roche Diagnostics GmbH, Germany), 250 U ml^−1^ benzonase (LubioScience GmbH, Switzerland), and 10 U ml^−1^ DNase I (Roche Diagnostics GmbH, Germany). Samples were incubated at 4°C for 1 hour and then centrifuged at 13,000xg at 4 °C for 15min. The supernatant was transferred to a protein LoBind tube (Eppendorf SE, Germany) and the protein concentration was quantified using a commercial bicinchoninic acid assay (Thermo Fisher Scientific, USA). An amount of 5μg of protein per sample was digested (Trypsin, Promega Corporation, USA) using an AssayMAP Bravo liquid handling system (Agilent Technologies, USA) running the autoSP3 protocol according to Müller et al.(*81*). After sample preparation, the remaining peptides were vacuum dried and stored at -20 °C until LC-MS/MS analysis.

#### MS method Orbitrap Astral Zoom

Samples were processed and measured in a block randomized order to prevent any technical bias that might occur during sample preparation or LC-MS/MS measurement (*82*).

Dried peptides were reconstituted (97.4% water, 2.5% hexafluoro-2-propanol and 0.1% trifluoroacetic acid) and 200ng were analysed using a Vanquish Neo UPLC system (Thermo Fisher Scientific, USA) directly connected to an Astral Zoom mass spectrometer for a total of 30 min per sample. Peptides were online desalted on a trapping cartridge (Acclaim PepMap300 C18, 5µm, 300Å wide pore; Thermo Fisher Scientific, USA) with a loading volume of 60µL using a 30 µL/min flow of 0.1% trifluoroacetic acid in water. The analytical multistep gradient (400 nL/min) was performed with an Aurora Ultimate XT analytical column (1.7 µm, 75 µm x 250 mm, IonOptics, Australia) using solvent A (0.1% formic acid in water) and solvent B (0.1% formic acid in acetonitrile). For 24.5 min the concentration of B linearly ramped from 2% to 30%, followed by a quick ramp to 80%.

Eluting peptides were analyzed in the mass spectrometer using data independent acquisition (DIA) mode. A full scan at 240k resolution (380-980 m/z, 500% AGC target, 5 ms maxIT) was followed by 300 DIA windows. The DIA acquisition covered a mass range of 380-980 m/z using windows of fixed width. Windows overlap by 2 m/z, the AGC target was 100% with a maxIT of 3 ms and spectra are recorded using Astral analyser. All laboratory technicians performing the measurement were blinded to the clinical data and group allocation.

#### Data analysis

Analysis of DIA RAW files was performed with Spectronaut (Biognosys, version 21.0.260703.94842, settings profile 2024-09-03_LFQ_MS) (*83*) in directDIA+ (Deep) library free mode. Default settings were applied with the following relevant parameters. Within DIA Analysis under Identification, the Precursor Qvalue Cutoff and Precursor PEP Cutoff were set to 0.01, and Protein Qvalue Cutoff (Run and Experiment) as well as Protein PEP Cutoff were set to 0.01; Run level protein scoring used the “Highest Scoring Observation (SN19 default)” rule. In Quantification, the Proteotypicity Filter was set to “Only Protein Group Specific”, the Protein LFQ Method was set to MaxLFQ and the Quantification window was set to “Not Synchronized (SN 17)”. In Post Analysis, “Use All MS Level Quantities” was set to True. The data were searched against the human reference proteome from UniProt (one protein sequence per gene; 20,659 entries, FASTA dated 29^th^ January 2026) and a contaminants FASTA (246 unique entries from 12^th^ of June 2025), and experimental conditions were included in the Spectronaut setup.

#### Data processing

The normalized data derived from Spectronaut was screened for gene name duplicates. Only the event with the highest number of identified precursors across all samples was kept. The dataset was then filtered for proteins that were quantified in ≥70% of the samples in at least one group × time point combination. Following log_2_ transformation, missing values were imputed separately within each group-by-time- point combination using the missForest package (*84*).

### Assessment of physical activity behavior

For subjective measurement of participants’ physical activity behavior, the Global Physical Activity Questionnaire (GPAQ) and the Godin Leisure-Time Exercise Questionnaire (GLTEQ) were used, respectively (*77,85*). The GLTEQ asks for weekly frequencies of strenuous (9 points), moderate (5 points), and mild (3 points) activities with all points being multiplied by the number of weekly occurrences. Reference score units are ≥ 24 (≙ active), 14 to 23 (≙ moderately active) and < 14 (≙ insufficiently active). The German GPAQ consists of 16 short-answer questions on sedentary behavior and physical activity participation in the three domains (1) activity at work, (2) travel to and from places, and (3) recreational activities and are categorized as moderate- or vigorous-intensity.

### Statistics

Data from a total of 75 persons with MS and 29 matched healthy subjects were available for statistical analyses. According to the flow diagram in Fig. 1, a total of 226 of the 254 possible samples were available for flow cytometry analysis. For three PBMC samples, no protein could be extracted, resulting in n = 223 for final proteomics analysis. Similarly, isolation of serum was not possible for three samples, resulting in n = 223 for sGFAP and n = 222 (one sample measurement failed) for sNfL analysis. An overview of the sample size per group × time point, including V͘O_2peak_, PPO, SII, and NLR, is given in Supplementary Table 5. Statistical analysis and visualization were performed in R (version 4.5.1). If not otherwise indicated, data wrangling and processing was mainly done using the dplyr (*86*) and tidyr (*87*) packages followed by visualization with ggplot2 (*88*).

### Baseline differences between persons with MS and healthy subjects

Data distribution was assessed for normality using the Shapiro-Wilk test and checked for linearity via quantile-quantile plots and histograms. In case of normal distribution, group differences were assessed using two-sided unpaired Student’s t-tests, otherwise the Wilcoxon-Mann-Whitney test was applied. The level of significance was set at p ≤ 0.05. Effect sizes were calculated using the effectsize package (*89*), with Cohen’s d (*d*) reported for parametric comparisons and the rank-biserial correlation (*r_rb_*) for nonparametric comparisons.

### Correlation analyses

Pearson’s correlation coefficient was used to assess associations between continuous variables, whereas Spearman’s rank correlation coefficient was used when at least one variable was ordinal (i.e., EDSS). Correlation coefficients are reported together with the corresponding two-sided p-values. For the exploratory correlation matrix comprising multiple pairwise comparisons (Fig. 5C), p- values were adjusted for multiple testing using the Benjamini-Hochberg procedure to control the false discovery rate (FDR). No adjustment for multiple testing was applied to the predefined correlation analyses presented as individual scatterplots (Supplementary Fig. S1C, S2).

### Effects of HIIT on physical fitness, immune cells, and targeted protein abundance

Group (intervention, passive control), time point (baseline, post- intervention, follow-up), and their interaction (group × time) were included as fixed effects, while subject ID was included as a random intercept and the baseline value as a fixed covariate (*90*). Linear mixed-effects models were fitted using the lmer function from the lmerTest package (*91*). The significance of the fixed effects (group, time point, and interaction) was then assessed using analysis of variance (ANOVA) via the built-in anova function from R stats. Results from targeted protein abundances derived from the proteomics dataset were adjusted for multiple testing using the Benjamini- Hochberg procedure (*92*) to control the FDR, while results derived from flow cytometry and physical fitness analyses were not corrected for multiple testing. Partial eta squared (*ƞp^2^*) effect sizes were reported for the main effects using the eta_squared function from the effectsize package (*89*). In case of significant results, pairwise comparisons of estimated marginal means between time points and/or groups were performed by applying the emmeans function from the emmeans package (*93*) and adjusted for multiple testing using the Bonferroni correction. Statistical significance was defined as an adjusted two-sided p-value of ≤ 0.05. Effect sizes (Cohen’s d) for significant pairwise comparisons (group and time) were calculated from the estimated marginal means using the residual standard deviation of the mixed model with the eff_size function from the emmeans package (*93*).

### Exercise-induced changes on the PBMC proteome

To identify proteins that are significantly altered between the intervention and passive control group, linear models were fitted to log_2_-transformed, normalized, and imputed protein intensities using the limma R package (*94*). To account for repeated measurements, within-subject correlation was estimated using the duplicateCorrelation function with subject ID as the blocking factor. The model included the group (intervention, passive control), the time point (baseline, post-intervention, follow-up), and their interaction (group × time) as fixed factors. Moderated t-statistics (*95*) were computed for each predefined contrast using the empirical Bayes approach implemented in the eBayes function with mean-variance trend correction and robust variance estimation. Resulting p-values were adjusted for multiple testing using the Benjamini-Hochberg procedure (*92*) to control the FDR, and statistical significance was defined as an adjusted two-sided p- value of ≤ 0.05.

### Transcription factor activity analysis

Transcription factor (TF) activities were inferred from the proteomics data using the DoRothEA human regulon (confidence levels A-C)(*37*) and the multivariate linear model implemented in the decoupleR package (*96*). Only regulons represented by at least 20 target proteins in the dataset were included in the analysis. TF activity scores were subsequently analyzed using linear models implemented in the limma R package (*94*). To account for repeated measurements, within-subject correlation was estimated using the duplicateCorrelation function with subject ID as the blocking factor. The model included the time point, group, and their interaction as fixed factors. Empirical Bayes moderation was applied using the eBayes function, and predefined contrasts were used to assess within-group temporal changes and group × time interactions. Resulting p-values were adjusted for multiple testing using the Benjamini-Hochberg procedure (*92*) to control the FDR, and statistical significance was defined as an adjusted two-sided p-value of ≤ 0.05.

### Coefficient of variation

Inter-individual variability was calculated as the coefficient of variation (CV) for each protein across all participants separated by group allocation (intervention, passive control) and time point (pre, post-intervention, follow-up) using non-log_2_ transformed, non-imputed, filtered data to avoid potential bias arising from imputation and data incompleteness. CVs were calculated as the ratio of the standard deviation σ to the mean μ.

## Data Availability

Raw data files of all samples processed for proteomics analysis are hosted on the PRoteomics IDEntifications Database (PRIDE) (URL will be made available upon manuscript acceptance). Raw data files of samples processed in the flow cytometry analysis are hosted on https://figshare.com (URL will be made available upon manuscript acceptance). All other data will be made available on request. All figures, except Fig. 1, were created using the indicated software (i.e., R packages).

## List of Supplementary Materials

Fig. S1. Exercise intervention-related data regarding implementation and efficacy.

Fig. S2. Pearson correlation analysis revealing a significant negative association between delta changes in V͘O2peak and the Neutrophil-to-Lymphocyte ratio within the intervention group (left), but not for the systemic immune-inflammation index (right).

Fig. S3. Gating strategy of flow cytometry analysis.

Fig. S4. Assessment of data robustness and potential confounders from PBMC proteomic analysis.

Fig. S5. Targeted analysis of proteins derived from the proteomics dataset that belong to the NF-κB (a) or STAT (b) signaling pathway.

Fig. S6. Cumulative and single expression levels of chemokine receptors and adhesion molecules on LFA-1α^low^ and LFA-1α^high^ T cells.

Fig. S7. Comparison of sGFAP (a) and sNfL (b) values between persons with MS and healthy subjects.

Table S1. Comparison of anthropometric and performance characteristics between persons with MS and matched healthy subjects divided by sex.

Table S2. Overview of flow cytometry and sNfL / sGFAP results including main effects and data per group and time point.

Table S3. Change in medication distribution per group and timepoint due to drop-out or missing assessment days.

Table S4. Overview of the block periodized high-intensity interval training intervention of 12 weeks.

Table S5. Overview of sample sizes by Group × Time point for each overarching analysis approach (i.e., proteomics, flow cytometry) or individual outcomes.

## Acknowledgements

We want to thank Franca Hegener for her help in planning and conducting the training sessions. We further thank Renata Panfilova for conducting sGFAP and sNfL analysis. Schematic Fig. 1 was created with BioRender (https://BioRender.com).

## Funding

P.Z., S.G.M., and C.A.O. acquired funding from the Deutsche Forschungsgemeinschaft (DFG, German Research Foundation) – Project number 594936650. S.P. received funding from the *Hans-Böckler-Stiftung*. T.W.C received funding from the *German Academic Exchange Service*. M.P. and S.G.M. acquired funding from the *German Multiple Sclerosis Foundation (DMSG)* Duesseldorf.

## CRediT authorship contribution statement

Conceptualization: P.Z., N.J., S.P., T.W.C.; Data Curation: S.P., T.W.C.; Formal Analysis: S.P.; Investigation: S.P., T.W.C., N.D., L.H., S.T., M.S., M.P., L.M., J.K.; P.Z.,

D.L.F.; Methodology: S.P., T.W.C., P.Z., N.J.; Resources: C.A.O., G.E-W., P.Z.;

Visualization: S.P.; Writing – Original Draft: S.P.; Writing – Review & Editing: T.W.C., L.H., S.T., N.D., N.J., D.W., G.E-W., S.A., M.S., P.Z., D.L.F., S.G.M., M.P., L.M., J.K.,

T.R., R.R., A.L.H., C.A.O., P.Z.; Supervision: P.Z., S.A., S.G.M.; Project Administration:

P.Z., S.P., T.W.C.; Funding Acquisition: S.P., T.W.C., P.Z., M.P.

### Competing interests

S.G.M. reports grants (DFG (German Research Foundation), BfR (German Federal Ministry of Food and Agriculture), DMSG (German Multiple Sclerosis Society), Hempel Foundation for Science, Art and Welfare, Ministry of Culture and Science of the state of North-Rhine-Westphalia, Heinrich-Heine University Düsseldorf), honoraria for lecturing, and travel expenses for attending meetings from Academy 2, Argenx, Alexion, Almirall, Amicus Therapeutics Germany, Bayer Health Care, Biogen, BioNtech, BMS, Celgene, Datamed, Demecan, Desitin, Diamed, Diaplan, DIU Dresden, DPmed, Gen Medicine and Healthcare products, Genzyme, Hexal AG, IGES, Impulze GmbH, Janssen Cilag, KW Medipoint, MedDay Pharmaceuticals, Medudy, Merck Serono, MICE, Mylan, Neuraxpharm, Neuropoint, Novartis, Novo Nordisk, ONO Pharma, Oxford PharmaGenesis, QuintilesIMS, Roche, Sanofi-Aventis, Springer Medizin Verlag, STADA, Chugai Pharma, Teva, UCB, Viatris, Wings for Life international and Xcenda. His research is funded by the German Ministry for Education and Research (BMBF), Bundesinstitut für Risikobewertung (BfR), Deutsche Forschungsgemeinschaft (DFG), Else Kröner Fresenius Foundation, Gemeinsamer Bundesausschuss (G-BA), German Academic Exchange Service, Hertie Foundation, Interdisciplinary Center for Clinical Studies (IZKF) Muenster, German Foundation Neurology.

M.P. received honoraria for lecturing and travel expenses for attending meetings from Alexion, ArgenX, Bayer Health Care, Biogen, Hexal, Merck Serono, Novartis, Roche, Sanofi-Aventis, Takeda and Teva. His research is funded by ArgenX, Biogen, Demecan, Hexal, Horizon Merck Serono, Novartis, Roche, Viatris, Takeda and Teva, all outside the scope of this work. His research is funded by the by the German Multiple

Sclerosis Foundation (DMSG), the B. Braun Foundation and the German Alzheimer Society.

L.M. reports honoraria for lecturing, consulting and travel expenses for attending meetings from Biogen, Merck, argenX, Roche, Neuraxpharm, Alexion and Novartis, all outside the scope of this work. His research is funded by the by the German Multiple Sclerosis Foundation (DMSG), the B.Braun Foundation and the Deutsche Forschungsgemeinschaft (DFG, German Research Foundation) – 493659010.

### Data and materials availability

Raw data files of all samples processed for proteomics analysis are hosted on the PRoteomics IDEntifications Database (PRIDE) (*URL will be made available upon manuscript acceptance*). Raw data files of samples processed in the flow cytometry analysis are hosted on https://figshare.com (*URL will be made available upon manuscript acceptance*). All other data will be made available on request. All figures, except Fig. 1, were created using the indicated software (i.e., R packages).

---

**Fig. S1.**
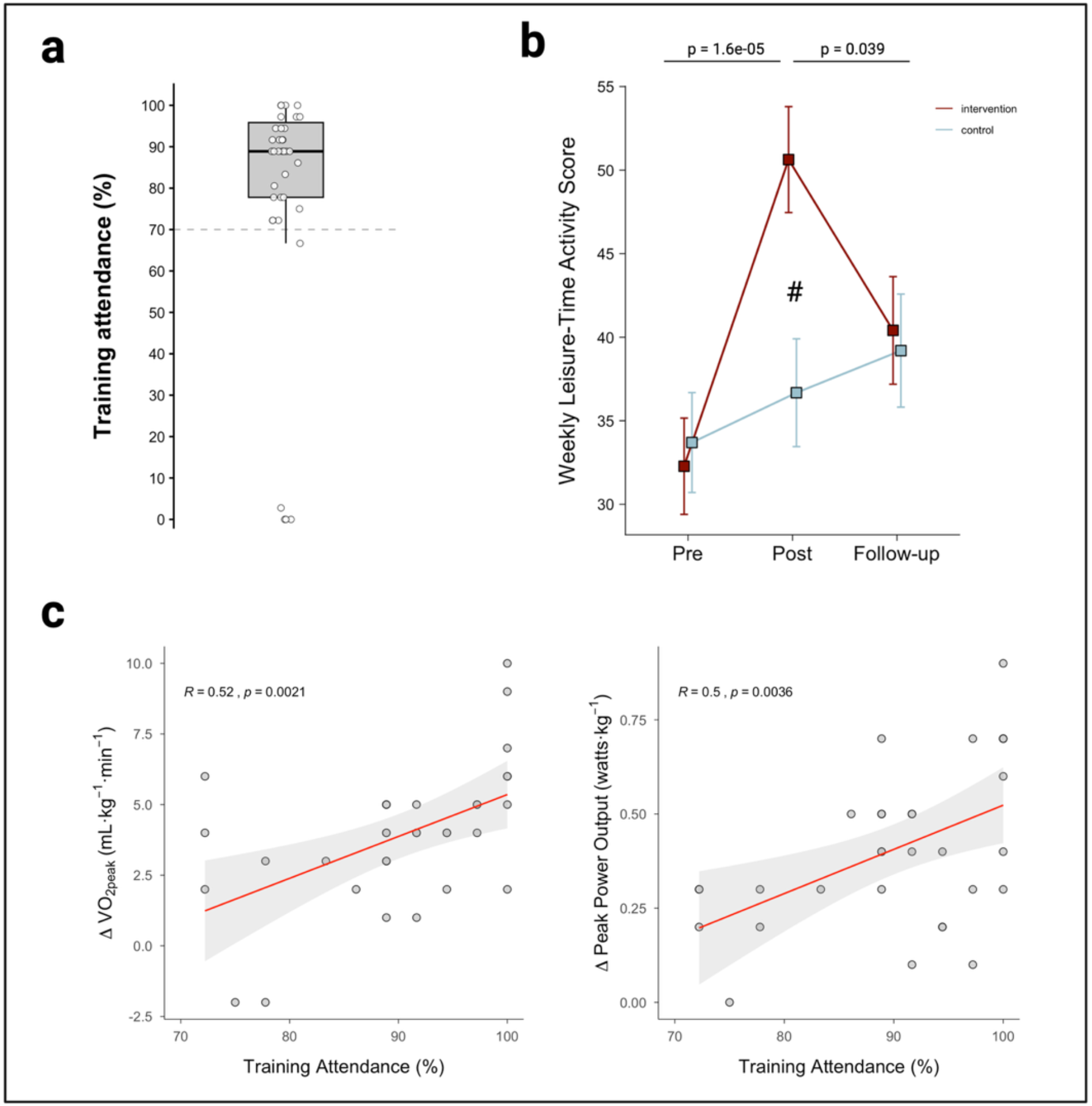
Exercise intervention-related data regarding implementation and efficacy. (a) Training attendance of participants in the intervention group. (b) Changes in weekly leisure-time activity over time in both groups using the Godin Leisure-Time Exercise Questionnaire. (c) Significant positive associations between the training attendance and the change in V͘O_2peak_ and peak power output in the intervention group. ^#^ indicate a significant group difference

**Fig. S2.**
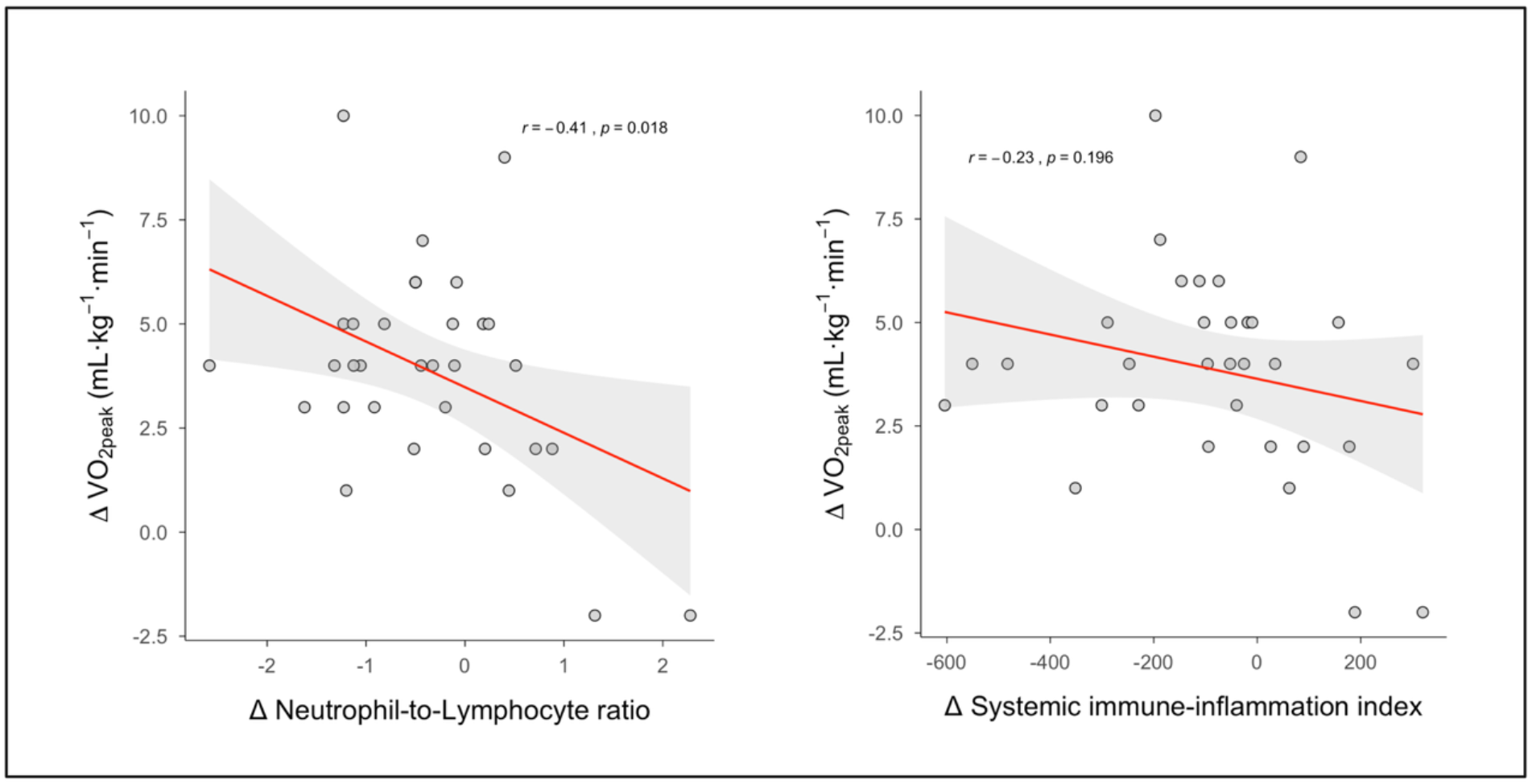
Pearson correlation analysis revealing a significant negative association between delta changes in V͘O_2peak_ and the Neutrophil-to-Lymphocyte ratio within the intervention group (left), but not for the systemic immune-inflammation index (right).

**Fig. S3.**
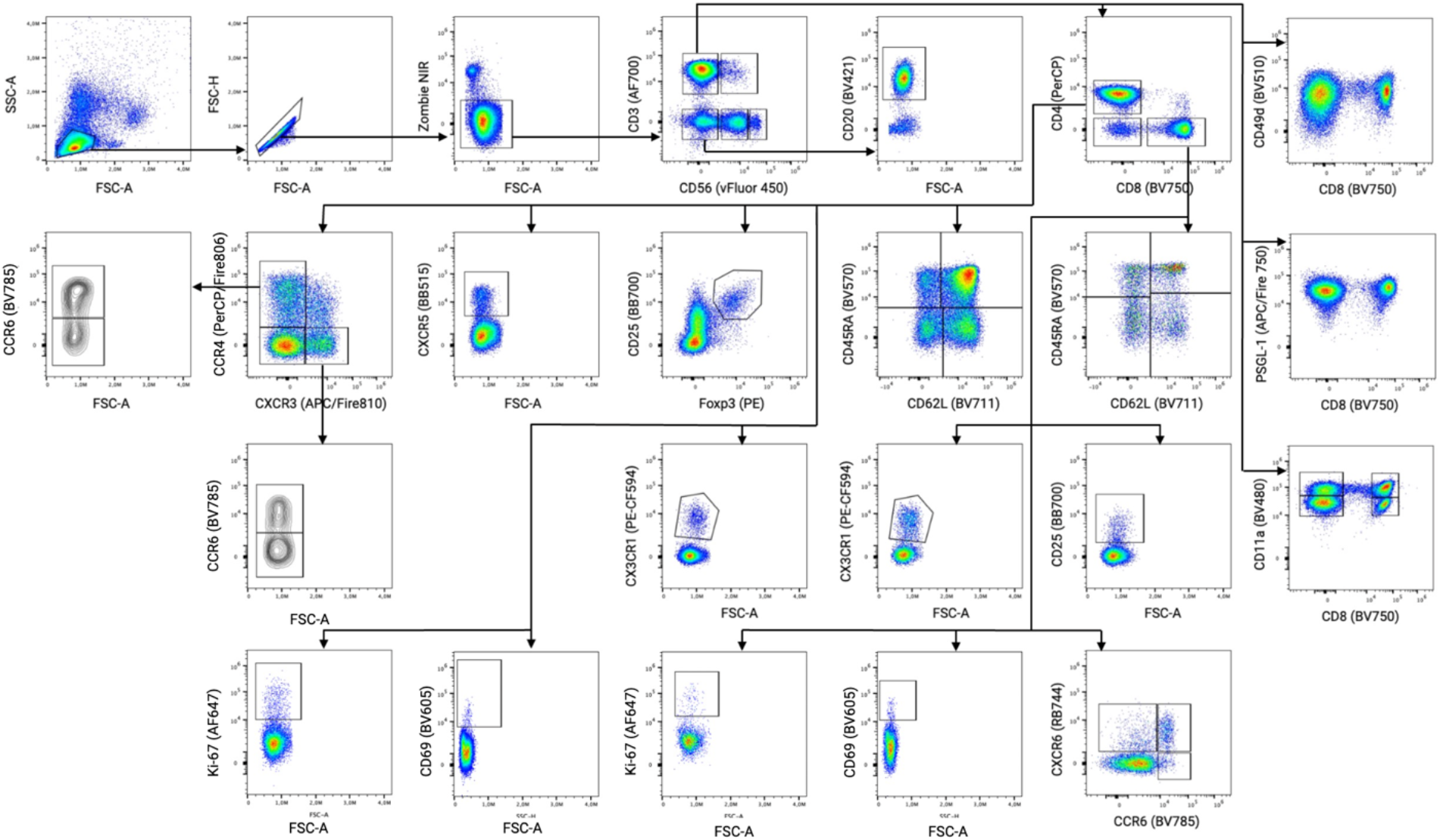
Gating strategy of flow cytometry analysis.

**Fig. S4.**
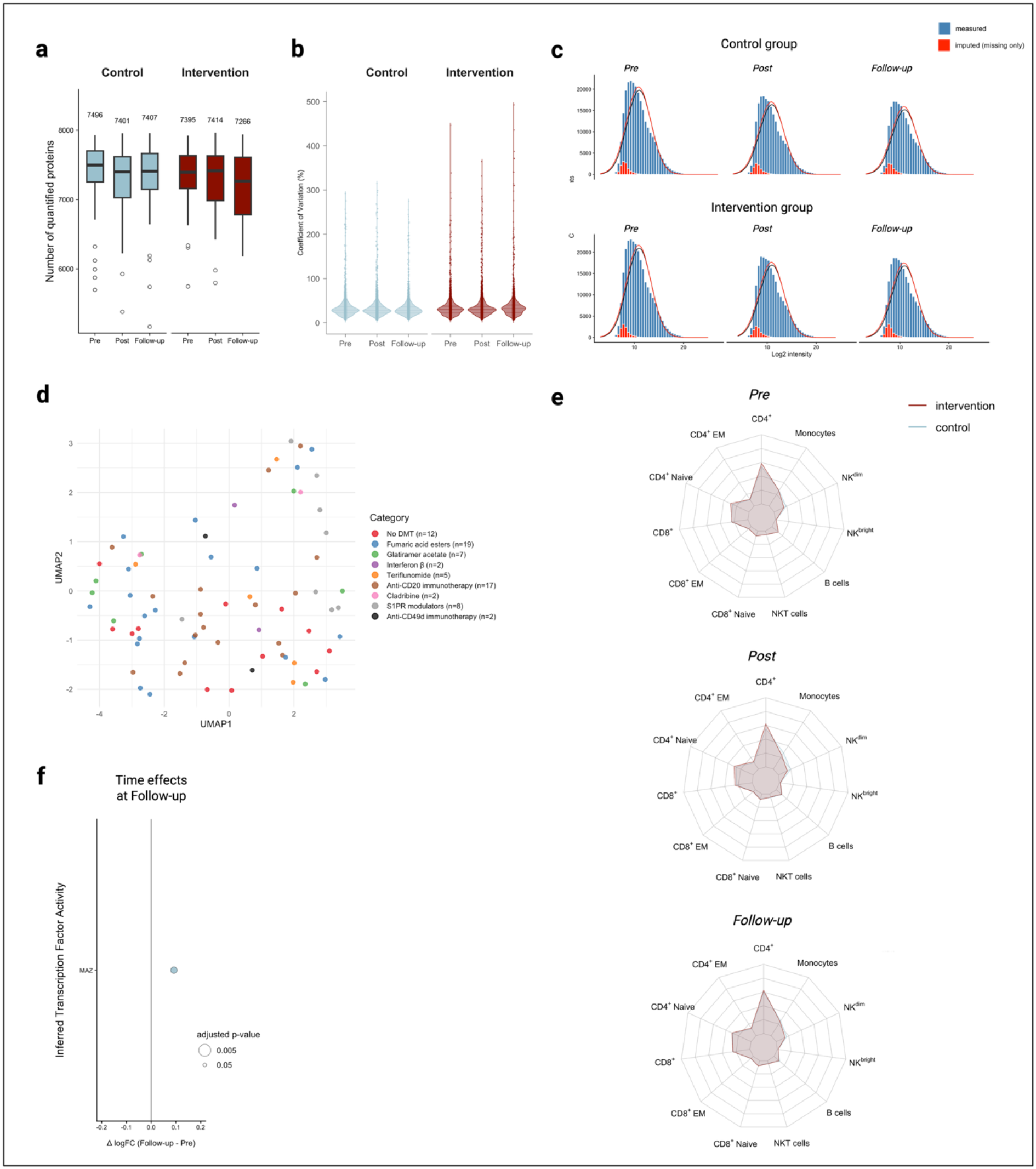
Assessment of data robustness and potential confounders from PBMC proteomic analysis. (a) Overview on the median number of protein quantification, (b) the coefficient of variation per protein, (c) and the imputation results that are plotted per group and time point. (d) Uniform Manifold Approximation and Projection (UMAP) clustering of baseline proteomic data per MS subject (n=74) with color-coding per DMT category. (e) Radar plots showing the distribution of major immune cell populations per group and time point. (f) Inferred changes in transcription factor activity in the control group from pre to follow- up based on the abundance of target proteins (n ≥ 20). DMT: disease-modifying therapy; S1PR: Sphingosine-1-phosphate receptor.

**Fig. S5.**
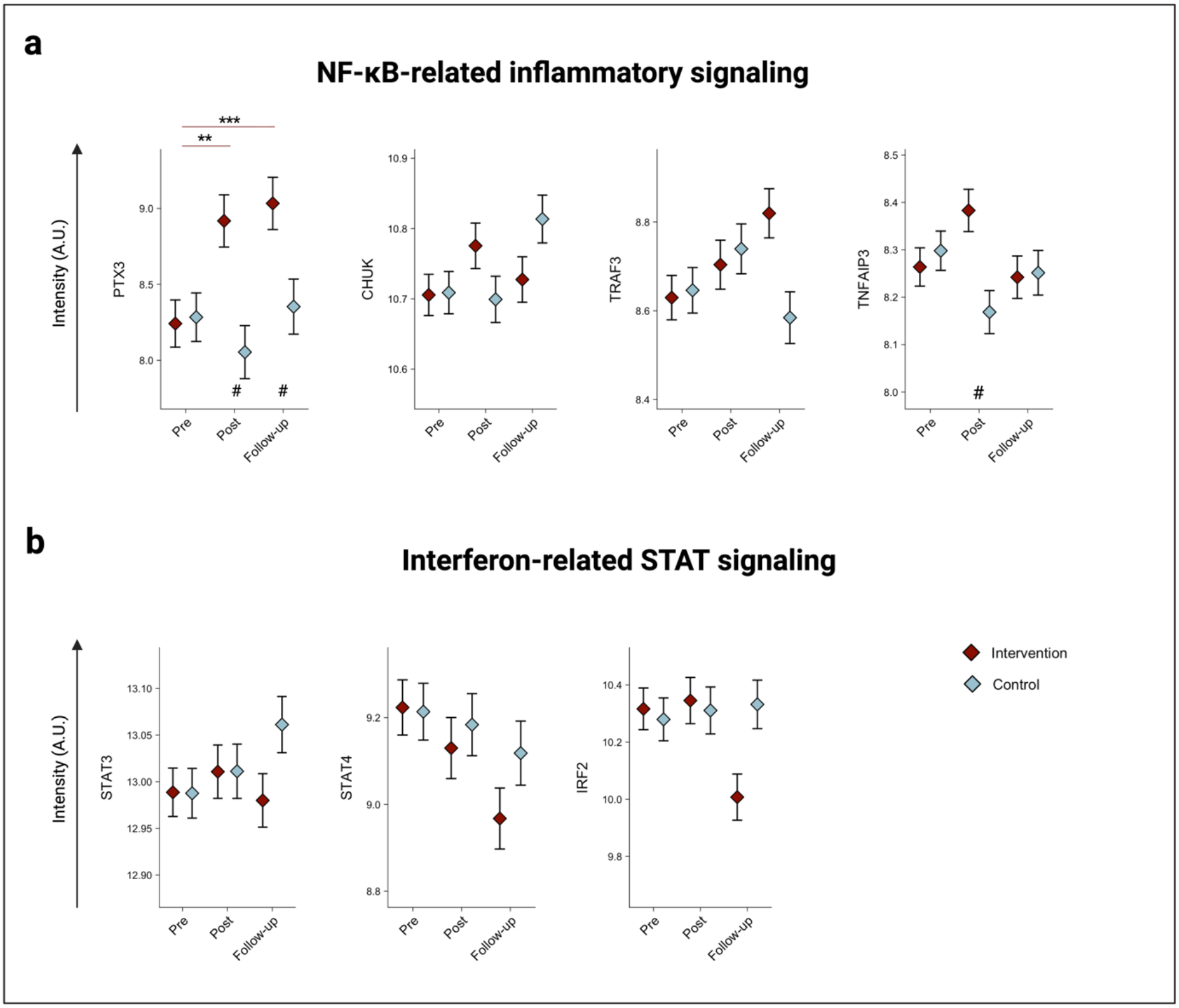
Targeted analysis of proteins derived from the proteomics dataset that belong to the NF-κB (a) or STAT (b) signaling pathway. *different from Pre (.05 ≥ p > .005). **different from Pre (.005 > p ≥ .001). ***different from Pre (p ≤ .001). ^#^ indicate a significant group difference.

**Fig. S6.**
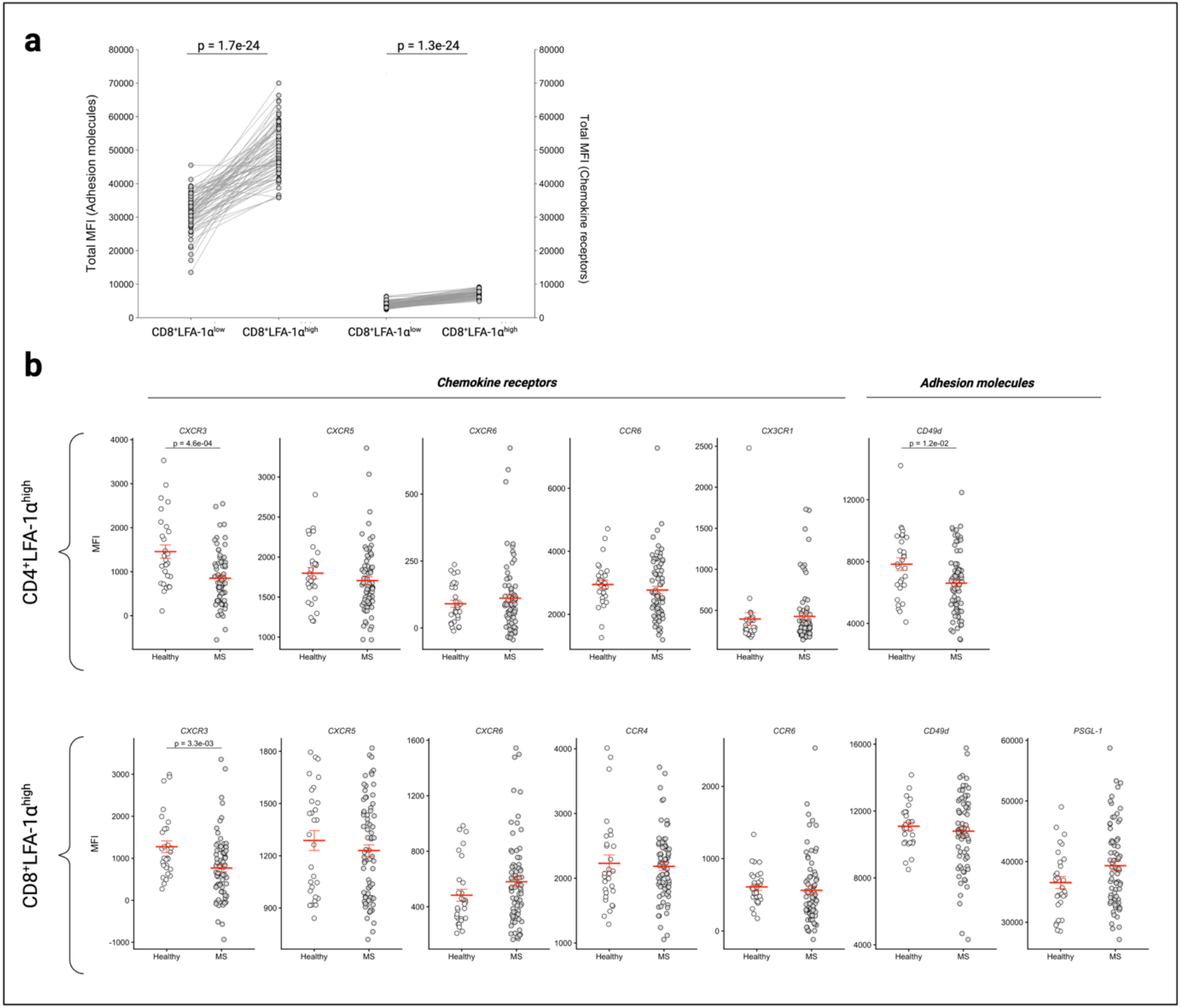
Cumulative and single expression levels of chemokine receptors and adhesion molecules on LFA-1α^low^ and LFA-1α^high^ T cells. (a) Comparison of the cumulative mean fluorescence intensity of adhesion molecules (i.e. PSGL-1 and CD49d) and chemokine receptors (i.e., CCR4, CCR6, CXCR3, CXCR5, CXCR6, and CX_3_CR1) between CD8^+^LFA-1α^low^ and a CD8^+^LFA-1α^high^ T cells. (b) Comparison of chemokine receptors and adhesion molecules within CD4^+^LFA-1α^high^ T cells (upper diagram) and CD8^+^LFA-1α^high^ T cells (lower diagram) between persons with MS and healthy subjects. MFI: Mean fluorescence intensity; MS: Multiple sclerosis.

**Fig. S7.**
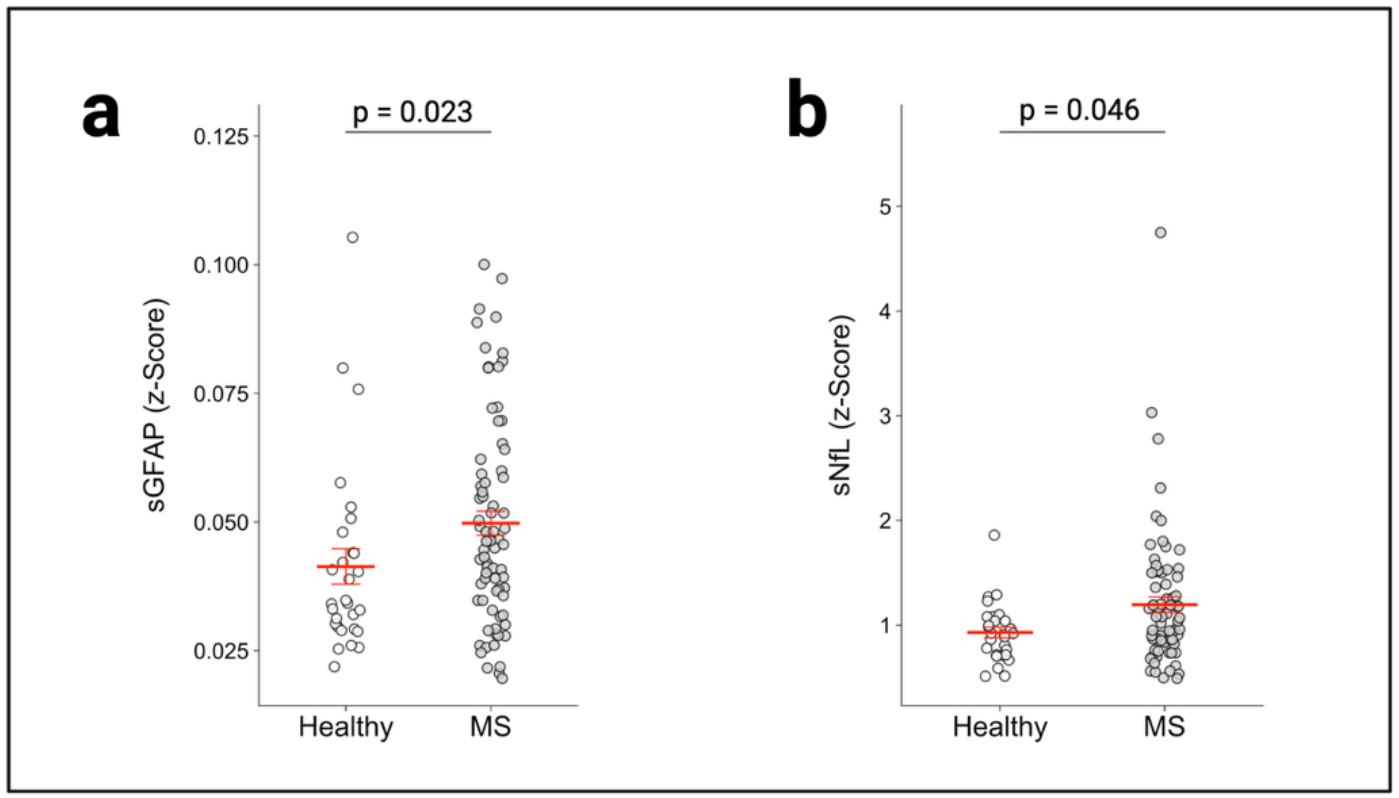
Comparison of sGFAP (a) and sNfL (b) values between persons with MS and healthy subjects. sGFAP: serum glial fibrillary acidic protein; sNfL: serum neurofilament light chain.

**Table S1.** Comparison of anthropometric and performance characteristics between persons with MS and matched healthy subjects divided by sex.

|  | Females (MS cohort, n=50) | Females (healthy cohort, n=19) | p value | Males (MS cohort n=25) | Males (healthy cohort, n=10) | p value |
|---|---|---|---|---|---|---|
| BMI (kg·m<sup>-2</sup>) | 25.9 ± 5.4 | 26.1 ± 5.9 | .89 | 26.2 ± 4 | 26.1 ± 3.4 | .93 |
| Age (years) | 43 ± 9.3 | 42.3 ± 11.3 | .80 | 40 ± 10.5 | 41.1 ± 12.3 | .80 |
| Smoker status (yes / no) | 6 / 44 | 4 / 15 |  | 4 / 21 | 1 / 9 |  |
| V̇O<sub>2peak</sub> (mL·kg<sup>-1</sup>·min<sup>-1</sup>) | 29.5 ± 5.4 | 31.3 ± 6.6 | .25 | 34.4 ± 7.8 | 39.2 ± 9.2 | .13 |
| PPO (W·kg<sup>-1</sup>) | 2.37 ± 0.5 | 2.65 ± 0.67 | .07 | 2.74 ± 0.74 | 3.49 ± 0.74 | .01 |

Data presented as mean ± SD. Two-sided unpaired t-tests were performed to compare characteristics between female (column 3) and male (column 6) subjects of the MS and healthy control cohort.
BMI: body mass index; EDSS: expanded disability status scale; f: female; m: male; PPO: peak power output; V̇O<sub>2peak</sub>: peak oxygen uptake.

**Table S2.**
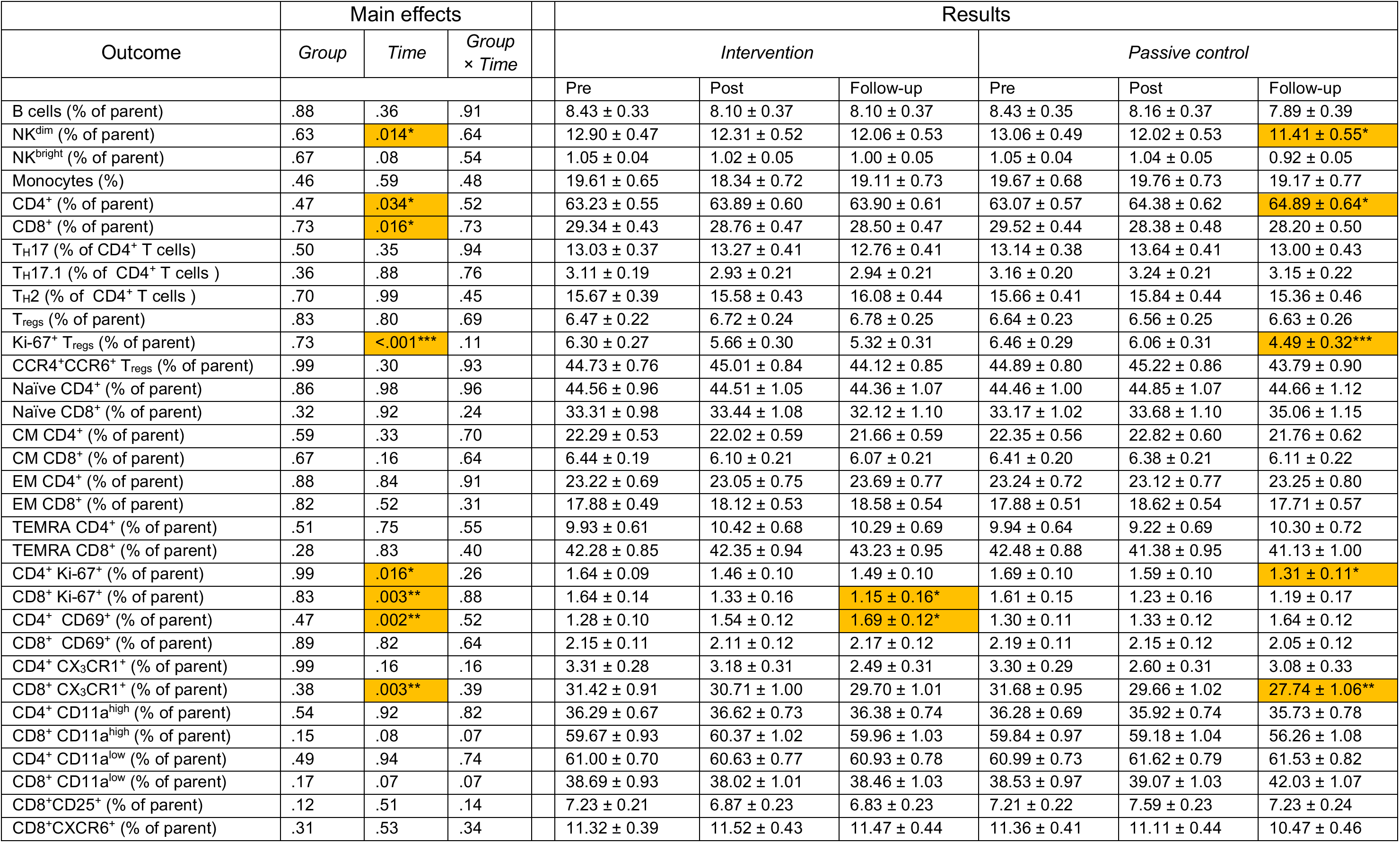
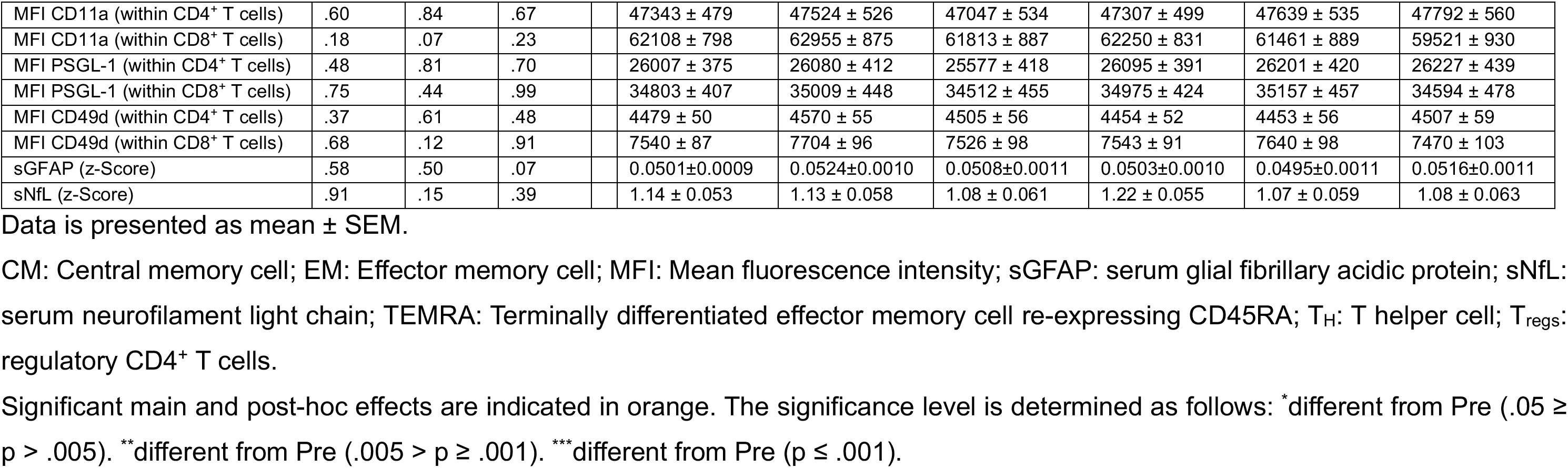
Overview of flow cytometry and sNfL / sGFAP results including main effects and data per group and time point.

|  | Main effects |  |  |  | Results |  |  |  |  |  |
|---|---|---|---|---|---|---|---|---|---|---|
| Outcome | *Group* | *Time* | *Group* × *Time* |  | *Intervention* |  |  | *Passive control* |  |  |
|  |  |  |  |  | Pre | Post | Follow-up | Pre | Post | Follow-up |
| B cells (% of parent) | .88 | .36 | .91 |  | 8.43 ± 0.33 | 8.10 ± 0.37 | 8.10 ± 0.37 | 8.43 ± 0.35 | 8.16 ± 0.37 | 7.89 ± 0.39 |
| NK<sup>dim</sup> (% of parent) | .63 | .014* | .64 |  | 12.90 ± 0.47 | 12.31 ± 0.52 | 12.06 ± 0.53 | 13.06 ± 0.49 | 12.02 ± 0.53 | 11.41 ± 0.55* |
| NK<sup>bright</sup> (% of parent) | .67 | .08 | .54 |  | 1.05 ± 0.04 | 1.02 ± 0.05 | 1.00 ± 0.05 | 1.05 ± 0.04 | 1.04 ± 0.05 | 0.92 ± 0.05 |
| Monocytes (%) | .46 | .59 | .48 |  | 19.61 ± 0.65 | 18.34 ± 0.72 | 19.11 ± 0.73 | 19.67 ± 0.68 | 19.76 ± 0.73 | 19.17 ± 0.77 |
| CD4<sup>+</sup> (% of parent) | .47 | .034* | .52 |  | 63.23 ± 0.55 | 63.89 ± 0.60 | 63.90 ± 0.61 | 63.07 ± 0.57 | 64.38 ± 0.62 | 64.89 ± 0.64* |
| CD8<sup>+</sup> (% of parent) | .73 | .016* | .73 |  | 29.34 ± 0.43 | 28.76 ± 0.47 | 28.50 ± 0.47 | 29.52 ± 0.44 | 28.38 ± 0.48 | 28.20 ± 0.50 |
| T<sub>H</sub>17 (% of CD4<sup>+</sup> T cells) | .50 | .35 | .94 |  | 13.03 ± 0.37 | 13.27 ± 0.41 | 12.76 ± 0.41 | 13.14 ± 0.38 | 13.64 ± 0.41 | 13.00 ± 0.43 |
| T<sub>H</sub>17.1 (% of CD4<sup>+</sup> T cells ) | .36 | .88 | .76 |  | 3.11 ± 0.19 | 2.93 ± 0.21 | 2.94 ± 0.21 | 3.16 ± 0.20 | 3.24 ± 0.21 | 3.15 ± 0.22 |
| T<sub>H</sub>2 (% of CD4<sup>+</sup> T cells ) | .70 | .99 | .45 |  | 15.67 ± 0.39 | 15.58 ± 0.43 | 16.08 ± 0.44 | 15.66 ± 0.41 | 15.84 ± 0.44 | 15.36 ± 0.46 |
| T<sub>regs</sub> (% of parent) | .83 | .80 | .69 |  | 6.47 ± 0.22 | 6.72 ± 0.24 | 6.78 ± 0.25 | 6.64 ± 0.23 | 6.56 ± 0.25 | 6.63 ± 0.26 |
| Ki-67<sup>+</sup> T<sub>regs</sub> (% of parent) | .73 | <.001*** | .11 |  | 6.30 ± 0.27 | 5.66 ± 0.30 | 5.32 ± 0.31 | 6.46 ± 0.29 | 6.06 ± 0.31 | 4.49 ± 0.32*** |
| CCR4<sup>+</sup>CCR6<sup>+</sup> T<sub>regs</sub> (% of parent) | .99 | .30 | .93 |  | 44.73 ± 0.76 | 45.01 ± 0.84 | 44.12 ± 0.85 | 44.89 ± 0.80 | 45.22 ± 0.86 | 43.79 ± 0.90 |
| Naïve CD4<sup>+</sup> (% of parent) | .86 | .98 | .96 |  | 44.56 ± 0.96 | 44.51 ± 1.05 | 44.36 ± 1.07 | 44.46 ± 1.00 | 44.85 ± 1.07 | 44.66 ± 1.12 |
| Naïve CD8<sup>+</sup> (% of parent) | .32 | .92 | .24 |  | 33.31 ± 0.98 | 33.44 ± 1.08 | 32.12 ± 1.10 | 33.17 ± 1.02 | 33.68 ± 1.10 | 35.06 ± 1.15 |
| CM CD4<sup>+</sup> (% of parent) | .59 | .33 | .70 |  | 22.29 ± 0.53 | 22.02 ± 0.59 | 21.66 ± 0.59 | 22.35 ± 0.56 | 22.82 ± 0.60 | 21.76 ± 0.62 |
| CM CD8<sup>+</sup> (% of parent) | .67 | .16 | .64 |  | 6.44 ± 0.19 | 6.10 ± 0.21 | 6.07 ± 0.21 | 6.41 ± 0.20 | 6.38 ± 0.21 | 6.11 ± 0.22 |
| EM CD4<sup>+</sup> (% of parent) | .88 | .84 | .91 |  | 23.22 ± 0.69 | 23.05 ± 0.75 | 23.69 ± 0.77 | 23.24 ± 0.72 | 23.12 ± 0.77 | 23.25 ± 0.80 |
| EM CD8<sup>+</sup> (% of parent) | .82 | .52 | .31 |  | 17.88 ± 0.49 | 18.12 ± 0.53 | 18.58 ± 0.54 | 17.88 ± 0.51 | 18.62 ± 0.54 | 17.71 ± 0.57 |
| TEMRA CD4<sup>+</sup> (% of parent) | .51 | .75 | .55 |  | 9.93 ± 0.61 | 10.42 ± 0.68 | 10.29 ± 0.69 | 9.94 ± 0.64 | 9.22 ± 0.69 | 10.30 ± 0.72 |
| TEMRA CD8<sup>+</sup> (% of parent) | .28 | .83 | .40 |  | 42.28 ± 0.85 | 42.35 ± 0.94 | 43.23 ± 0.95 | 42.48 ± 0.88 | 41.38 ± 0.95 | 41.13 ± 1.00 |
| CD4<sup>+</sup> Ki-67<sup>+</sup> (% of parent) | .99 | .016* | .26 |  | 1.64 ± 0.09 | 1.46 ± 0.10 | 1.49 ± 0.10 | 1.69 ± 0.10 | 1.59 ± 0.10 | 1.31 ± 0.11* |
| CD8<sup>+</sup> Ki-67<sup>+</sup> (% of parent) | .83 | .003** | .88 |  | 1.64 ± 0.14 | 1.33 ± 0.16 | 1.15 ± 0.16* | 1.61 ± 0.15 | 1.23 ± 0.16 | 1.19 ± 0.17 |
| CD4<sup>+</sup> CD69<sup>+</sup> (% of parent) | .47 | .002** | .52 |  | 1.28 ± 0.10 | 1.54 ± 0.12 | 1.69 ± 0.12* | 1.30 ± 0.11 | 1.33 ± 0.12 | 1.64 ± 0.12 |
| CD8<sup>+</sup> CD69<sup>+</sup> (% of parent) | .89 | .82 | .64 |  | 2.15 ± 0.11 | 2.11 ± 0.12 | 2.17 ± 0.12 | 2.19 ± 0.11 | 2.15 ± 0.12 | 2.05 ± 0.12 |
| CD4<sup>+</sup> CX<sub>3</sub>CR1<sup>+</sup> (% of parent) | .99 | .16 | .16 |  | 3.31 ± 0.28 | 3.18 ± 0.31 | 2.49 ± 0.31 | 3.30 ± 0.29 | 2.60 ± 0.31 | 3.08 ± 0.33 |
| CD8<sup>+</sup> CX<sub>3</sub>CR1<sup>+</sup> (% of parent) | .38 | .003** | .39 |  | 31.42 ± 0.91 | 30.71 ± 1.00 | 29.70 ± 1.01 | 31.68 ± 0.95 | 29.66 ± 1.02 | 27.74 ± 1.06** |
| CD4<sup>+</sup> CD11a<sup>high</sup> (% of parent) | .54 | .92 | .82 |  | 36.29 ± 0.67 | 36.62 ± 0.73 | 36.38 ± 0.74 | 36.28 ± 0.69 | 35.92 ± 0.74 | 35.73 ± 0.78 |
| CD8<sup>+</sup> CD11a<sup>high</sup> (% of parent) | .15 | .08 | .07 |  | 59.67 ± 0.93 | 60.37 ± 1.02 | 59.96 ± 1.03 | 59.84 ± 0.97 | 59.18 ± 1.04 | 56.26 ± 1.08 |
| CD4<sup>+</sup> CD11a<sup>low</sup> (% of parent) | .49 | .94 | .74 |  | 61.00 ± 0.70 | 60.63 ± 0.77 | 60.93 ± 0.78 | 60.99 ± 0.73 | 61.62 ± 0.79 | 61.53 ± 0.82 |
| CD8<sup>+</sup> CD11a<sup>low</sup> (% of parent) | .17 | .07 | .07 |  | 38.69 ± 0.93 | 38.02 ± 1.01 | 38.46 ± 1.03 | 38.53 ± 0.97 | 39.07 ± 1.03 | 42.03 ± 1.07 |
| CD8<sup>+</sup>CD25<sup>+</sup> (% of parent) | .12 | .51 | .14 |  | 7.23 ± 0.21 | 6.87 ± 0.23 | 6.83 ± 0.23 | 7.21 ± 0.22 | 7.59 ± 0.23 | 7.23 ± 0.24 |
| CD8<sup>+</sup>CXCR6<sup>+</sup> (% of parent) | .31 | .53 | .34 |  | 11.32 ± 0.39 | 11.52 ± 0.43 | 11.47 ± 0.44 | 11.36 ± 0.41 | 11.11 ± 0.44 | 10.47 ± 0.46 |
| MFI CD11a (within CD4<sup>+</sup> T cells) | .60 | .84 | .67 |  | 47343 ± 479 | 47524 ± 526 | 47047 ± 534 | 47307 ± 499 | 47639 ± 535 | 47792 ± 560 |
| MFI CD11a (within CD8<sup>+</sup> T cells) | .18 | .07 | .23 |  | 62108 ± 798 | 62955 ± 875 | 61813 ± 887 | 62250 ± 831 | 61461 ± 889 | 59521 ± 930 |
| MFI PSGL-1 (within CD4<sup>+</sup> T cells) | .48 | .81 | .70 |  | 26007 ± 375 | 26080 ± 412 | 25577 ± 418 | 26095 ± 391 | 26201 ± 420 | 26227 ± 439 |
| MFI PSGL-1 (within CD8<sup>+</sup> T cells) | .75 | .44 | .99 |  | 34803 ± 407 | 35009 ± 448 | 34512 ± 455 | 34975 ± 424 | 35157 ± 457 | 34594 ± 478 |
| MFI CD49d (within CD4<sup>+</sup> T cells) | .37 | .61 | .48 |  | 4479 ± 50 | 4570 ± 55 | 4505 ± 56 | 4454 ± 52 | 4453 ± 56 | 4507 ± 59 |
| MFI CD49d (within CD8<sup>+</sup> T cells) | .68 | .12 | .91 |  | 7540 ± 87 | 7704 ± 96 | 7526 ± 98 | 7543 ± 91 | 7640 ± 98 | 7470 ± 103 |
| sGFAP (z-Score) | .58 | .50 | .07 |  | 0.0501±0.0009 | 0.0524±0.0010 | 0.0508±0.0011 | 0.0503±0.0010 | 0.0495±0.0011 | 0.0516±0.0011 |
| sNfL (z-Score) | .91 | .15 | .39 |  | 1.14 ± 0.053 | 1.13 ± 0.058 | 1.08 ± 0.061 | 1.22 ± 0.055 | 1.07 ± 0.059 | 1.08 ± 0.063 |

Data is presented as mean ± SEM.
CM: Central memory cell; EM: Effector memory cell; MFI: Mean fluorescence intensity; sGFAP: serum glial fibrillary acidic protein; sNfL: serum neurofilament light chain; TEMRA: Terminally differentiated effector memory cell re-expressing CD45RA; T<sub>H</sub>: T helper cell; T<sub>regs</sub>: regulatory CD4<sup>+</sup> T cells.
Significant main and post-hoc effects are indicated in orange. The significance level is determined as follows: <sup>\*</sup>different from Pre (.05 ≥ p > .005). <sup>\*\*</sup>different from Pre (.005 > p ≥ .001). <sup>\*\*\*</sup>different from Pre (p ≤ .001).

**Table S3.**
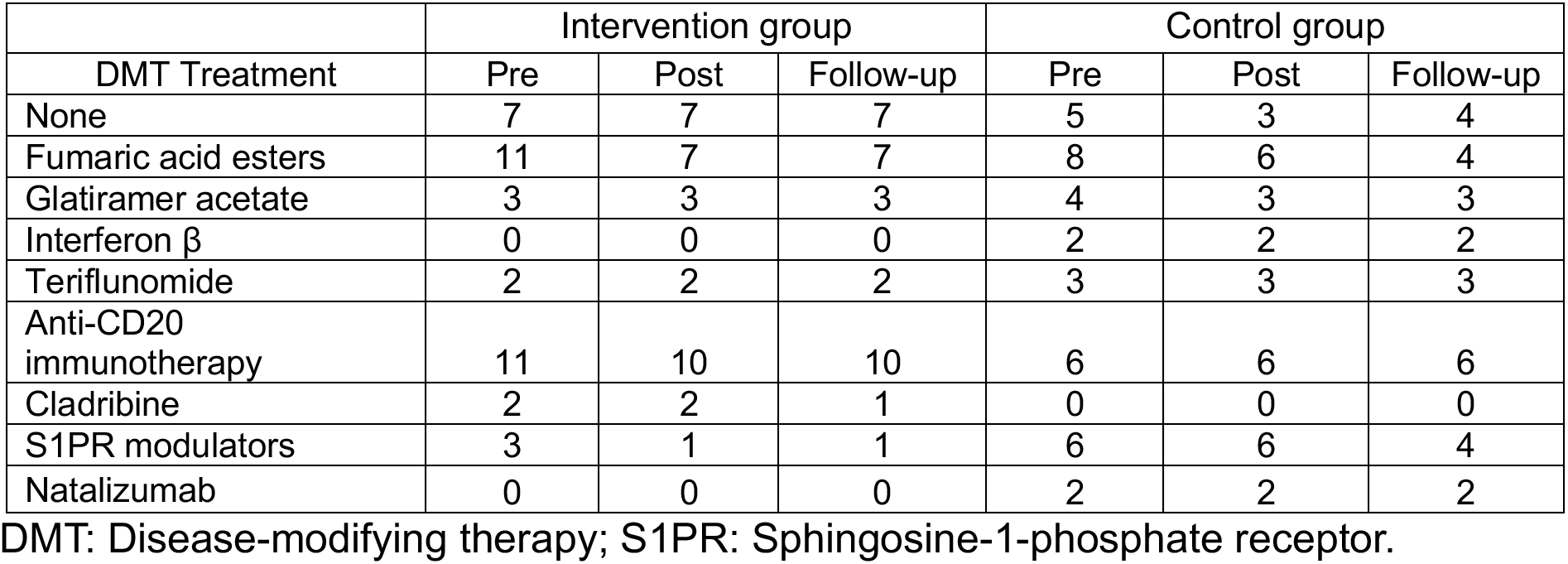
Change in medication distribution per group and timepoint due to drop-out or missing assessment days.

**Table S4.** Overview of the block periodized high-intensity interval training intervention of 12 weeks. During each block, participants exercised three times per week. During intervention week 6, another cardiopulmonary exercise test was conducted in order to update peak power output values that are used for controlling exercise intensity.

|  | Week 1-3 | Week 4-6 | Week 7-9 | Week 10-12 |
|---|---|---|---|---|
| Session duration | 38.5 min | 38.5 min | 38.5 min | 38.5 min |
| Intervals | 7x1.5 min | 7x1.5 min | 7x1.5 min | 7x1.5 min |
| Active recovery phase | 3 min | 3 min | 3 min | 3 min |
| Interval Intensity | 70-75% PPO | 80-85% PPO | 85-90% PPO | >90% PPO |
| Recovery phase intensity | 20-25% PPO | 20-25% PPO | 20-25% PPO | 20-25% PPO |
| Warm-up/cool-down | 5 min/5 min | 5 min/5 min | 5 min/5 min | 5 min/5 min |

PPO: Peak Power Output in Watts.

**Table S5.** Overview of sample sizes by Group × Time point for each overarching analysis approach (i.e., proteomics, flow cytometry) or individual outcomes.

|  | Overarching analysis approach |  | Individual outcomes |  |  |  |  |  |
|---|---|---|---|---|---|---|---|---|
| Group × Time point | PBMC Proteomics | Flow cytometry | sGFAP | sNfL | V̇O<sub>2peak</sub> | PPO | SII | NLR |
| Intervention (Baseline) | 38 | 39 | 39 | 38 | 39 | 39 | 39 | 39 |
| Intervention (Post) | 31 | 32 | 32 | 32 | 32 | 32 | 32 | 32 |
| Intervention (Follow-up) | 31 | 31 | 29 | 29 | 31 | 31 | 31 | 31 |
| Control (Baseline) | 36 | 36 | 36 | 36 | 36 | 36 | 36 | 36 |
| Control (Post) | 30 | 31 | 31 | 31 | 29 | 29 | 31 | 31 |
| Control (Follow-up) | 28 | 28 | 27 | 27 | 26 | 26 | 27 | 27 |
| Healthy (Baseline) | 29 | 29 | 29 | 29 | 29 | 29 | 28 | 29 |
| ∑ | **223** | **226** | **223** | **222** | **222** | **222** | **224** | **225** |

GFAP: serum glial fibrillary acidic protein; NLR: Neutrophil-to-Lymphocyte Ratio; PBMC: Peripheral blood mononuclear cells; PPO: peak power output; sNfL: serum neurofilament light chain; SII: Systemic Immune-inflammation Index; V̇O<sub>2peak</sub>: peak oxygen uptake.

## Notes

### Clinical Trial

DRKS00033055

### Author Declarations

The the ethics committee of the Leibniz Institute at TU Dortmund University gave ethical approval for this work

